# Developmental origins of the Autism Spectrum Disorder in the Middle East and North Africa Region: A Systematic Review and Meta-Analysis of adjusted risk factors

**DOI:** 10.1101/2024.08.27.24312654

**Authors:** Aishat F. Akomolafe, Fathima R. Mahmood, Bushra M. Abdallah, Amgad M. Elshoeibi, Elhassan Mahmoud, Aisha Abdulla Al-Khulaifi, Nour Darwish, Yara Dweidri, Duaa Yousif, Hafsa Khalid, Majed Al-Theyab, Muhammad Waqar Azeem, Durre Shahwar, Madeeha Kamal, Majid Alabdulla, Salma M. Khaled, Tawanda Chivese

**Author notes:** Corresponding and guarantor author: Tawanda Chivese, Department of Population Medicine, College of Medicine, QU Health, Qatar University, Doha, Qatar P O BOX 2713 Doha, Qatar.

## Abstract

**Background and Objectives:** The etiology of autism spectrum disorder (ASD) is poorly understood, with sparse data from the Middle East and North Africa (MENA) region, which has a unique climate and socio-epidemiological setting. This research investigated the developmental (early life) risk factors associated with ASD in the MENA region.

**Methods:** In this systematic review and meta-analysis, we searched for observational studies, which carried out adjusted analyses for ASD risk factors in the MENA region, in PubMed, Embase, Scopus, and CINAHL databases. We analyzed associations between ASD and factors related to conception, inheritance, maternal morbidity during pregnancy and adverse pregnancy outcomes. After study quality assessment, meta-analyses for each risk factor were carried out using the bias-adjusted inverse variance heterogeneity model. Heterogeneity was assessed using I^2^ and publication bias using Doi and funnel plots.

**Results:** The systematic review included 19 case control studies from eight countries within the MENA region. In overall synthesis, male sex (OR=3.27, 95% CI: 2.39-4.48, I2 = 40.9%), family history of ASD (OR=2.98, 95% CI: 0.51-17.31, I2 = 85.0%), and consanguinity (OR= 1.77, 95% CI: 1.38-2.28, I2 = 57.1%) were associated with ASD. Although with limited studies, a review of the literature showed some possible associations between ASD and gestational diabetes, gestational hypertension, macrosomia, NICU admission, respiratory distress syndrome, cesarean delivery, gestational age, and older maternal age.

**Conclusion:** These findings confirm the association between male sex, family history of ASD and consanguinity with ASD, potentially suggesting some inherited mechanism in the etiology of ASD in the MENA region. Some maternal co-morbidities during pregnancy and adverse pregnancy outcomes may contribute to ASD risk in the MENA region, although more studies are needed in the region.

**Registration:** The protocol for this systematic review and meta-analysis is registered on the International Prospective Register of Systematic Reviews (PROSPERO) with registration ID CRD42024499837.

## 1. Introduction

Autism spectrum disorder (ASD) is a developmental condition, characterized by impairments in social interactions, communication skills, and behavior, typically diagnosed in early childhood (1). ASD lies on a spectrum, varying in severity and presenting symptoms, with first symptoms usually emerging by the second year of life, classifying it as a developmental disorder (2). ASD imposes significant health and social burdens (3). The disorder also carries substantial economic repercussions for society and affected families due to the specialized care and significant financial resources required (4). Moreover, individuals with ASD often face high rates of mental health issues, stigma, and, in many countries, encounter challenges in employment (5).

Data from the World Health Organization (WHO), estimates autism to approximately 1 in 100 children worldwide (6). This figure represents an average value as reported rates vary significantly between countries, populations and regions. The prevalence remains unknown in many low-and middle-income countries (LMIC), a grouping which contains most countries in the Middle East and North Africa (MENA) region (6). In the Arab and Gulf countries, a systematic review in 2014 reported that the prevalence for ASD ranged from 1.4 to 29 per 10,000 persons in the region indicating the possibility of higher prevalence rates in this region compared to global rates (7). Moreover, this study identified advanced maternal and paternal age, cesarean section, prenatal complications, and suboptimal breastfeeding as potential contributors to autism and suggested that further investigation is necessary to enhance understanding of the factors contributing to the prevalence of autism in this region.

The risk factors for ASD are poorly understood, with many factors having been researched. These factors are best understood through the concept of the Developmental Origins of Health and Disease (DOHaD) which asserts that an individual’s health and disease profile is heavily influenced by early life events and modified by later risk factors (8). DOHaD theory highlights critical developmental periods, particularly during pregnancy and the first 1000 days, where factors may influence the risk for ASD. The developmental factors include inheritance-related factors such as family history of ASD, child sex, consanguinity, and certain genetic conditions like Down Syndrome (9–11) (12). Other developmental factors include conception-related and pregnancy-related factors such as maternal and paternal age, maternal comorbidities in pregnancy, and adverse pregnancy outcomes such as low birthweight and caesarian delivery (13, 14). Early childhood exposures such as birth weight, breastfeeding, infections, chronic diseases, undernutrition, and vaccination status have also been studied (13, 14). Consanguinity, referring to marriage or reproduction between close relatives, is prevalent in many MENA countries, and has also been linked to ASD(12, 15). However, studies have shown conflicting results and debate is ongoing about the developmental causes of ASD globally, and especially in the MENA region.

Despite ongoing research, consensus on the potential causes and risk factors of ASD remains elusive, with a notable scarcity of meta-analytical studies investigating this important topic in the MENA region. Therefore, the present systematic review and meta-analysis aims to identify potential risk factors for ASD in the MENA region. It fills an important gap in the literature on the contributing factors for the development of ASD and informs the future development of preventive strategies and targeted interventions. The primary objective was to identify and quantify the association between potential risk factors and ASD in the MENA.

## 2. Methods

### 2.1. Study Design and Protocol Registration

This systematic review and meta-analysis of observational studies adheres to the Preferred Reporting Items for Systematic Reviews and Meta-Analyses (PRISMA) 2020 guidelines (Supplementary Table S1) (16). The study protocol is registered in the International Prospective Register of Systematic Reviews (PROSPERO) with the registration number CRD42024499837.

### 2.2. Data sources

We conducted searches across multiple databases, including PubMed, Embase, Scopus, and CINAHL, without language or date restrictions, with the last search on 20 January 2024. Additionally, we screened the references of included studies to identify any additional relevant research. In cases where a full-text report was unavailable, authors were contacted directly to request for the study reports.

### 2.3. Search Methods

On the 20^th^ of January 2024, we crafted our search strategy for the paper by utilizing PubMed’s Medical Subject Headings (MeSH) terms, including "Autism Spectrum Disorder"[Mesh], "Autistic Disorder"[Mesh], and "Asperger Syndrome"[Mesh]. Additionally, we incorporated title and abstract terms for ASD and its synonymous expressions, along with relevant terms for the MENA region, which encompassed a comprehensive list of countries within this geographical area. Using the Polyglot translator, we adapted the developed search strategy for use in Embase, Scopus, Medline (EBSCO), and CINHAL databases (17). During the initial search, we imposed no restrictions regarding language or publication date. Supplementary Tables S2 – S5 contain the complete search strategy utilized for each database.

### 2.4. Procedure for Selection of Studies

Study records identified from each database search were transferred to EndNote 20 for duplicate identification and removal. They were then uploaded to the Rayyan Systematic Review Management platform for further duplicate removal and for title and abstract screening (18). Three pairs of independent investigators conducted manual assessments of the titles and abstracts for each article. In instances of disagreement between a pair, a third independent investigator was consulted for a final decision. Abstracts in languages other than English were translated using Google Translate and then assessed for eligibility. The same investigators conducted full-text screening for any articles deemed eligible during the initial title and abstract screening phase. Throughout the entire screening process, blinding was implemented.

### 2.5. Eligibility Criteria

The eligibility criteria for considering studies for this review included any type of observational studies that assessed the risk factors for ASD in the MENA region. Inclusion criteria encompassed all published and unpublished observational studies that focused on the population in the MENA region and reported risk factors for ASD in children. Studies were included only if they reported adjusted associations between investigated risk factors and ASD. Studies were excluded if they included populations which resided outside the MENA, if they were duplicate publications (with the least informative duplicate being excluded), if they did not contain primary data, and if they were case series, case reports, narrative reviews, letters to the editor, and opinions.

### 2.6. Outcomes

The primary outcome of interest was the development of ASD, diagnosed according to DSM-V diagnostic criteria. The DSM-V criteria evaluates autism based on 1.) Persistent deficits in social communication and social interaction across multiple contexts. 2.) Restricted, repetitive patterns of behavior, interests, or activities. 3.) Symptoms must be present in the early developmental period. 4.) Symptoms cause clinically significant impairment. 5.) These disturbances are not better explained by intellectual disability or global developmental delay.

### 2.7. Data Extraction

Two pairs of authors independently extracted data from the selected studies into a standardized data extraction form using Microsoft Office Excel. If discrepancies arose, the two researchers compared their results and discussed them to address the discrepancies. The extracted data included study characteristics such as the authors, year of publication, country where the study was conducted, study design, study period, sample size, sampling method, the risk factors described in the study, the type of analysis carried out and the covariates adjusted for. For each potential risk factor of interest, we extracted the adjusted effect size along with its 95% CIs, and p-values. We were interested in the following risk factors described in the literature: conception-related such as maternal and paternal age, inheritance-related factors, pregnancy-related morbidities, and adverse pregnancy outcomes as risk factors of ASD.

### 2.8. Assessment of the Quality of Included Studies

The assessment of the quality of the included studies was performed by two pairs of independent investigators using the Methodological Standard for Epidemiological Research (MASTER) scale (19). The MASTER scale consists of seven methodological standards with a total of thirty-six safeguards. The seven standards include: equal recruitment, equal retention, equal ascertainment, equal implementation, equal prognosis, sufficient analysis, and temporal precedence. Any disagreement between the two investigators in the assessment of the quality of the studies was resolved through discussion.

### 2.9. Data Synthesis

Data that could not be included in the meta-analysis were analyzed descriptively. For risk factors with three or less studies, only a systematic review of the literature was carried out. For the meta-analysis, we used the bias adjusted inverse variance heterogeneity (quality effects) model to perform bias-adjusted synthesis of the effect sizes for each risk factor CIs. For one study on sex as a risk factor (20), the inverse of the OR and 95% CI was used as the reference group used in the study was males instead of females. This model utilizes quality scores to rank studies based on their methodological rigor, adjusting the weights of each study accordingly to account for variability in quality (21, 22). To assess heterogeneity among the studies for each risk factor, we used the I² statistic and Cochran’s Q p-value (23). Publication bias and small study effects were evaluated using funnel plots, the Doi plot and LFK index (24). Significant heterogeneity was defined as a Cochran Q test p-value <0.05 or I² >50%. Standard funnel plots and Egger’s regression test p-value were done to validate the performance of the Doi plots and LFK index (25). Sensitivity was carried out using a leave-one-out analysis. All analyses were conducted using Stata 17.0.

## 3. Results

### 3.1. Search Results

The search identified a total of 3072 study records. Of these, 1630 studies were excluded as duplicates. Following title and abstract screening, 223 records underwent full-text screening. Five full texts could not be retrieved and were hence excluded. Lastly, 196 studies were excluded for the reasons listed in the below PRISMA Flowchart (Fig. 1) and in Supplementary Table S6. The final number of studies that were included in the systematic review was 19. Only 1 of these studies (26) was not included in a meta-analysis due to the different cutoff point used for paternal age to assess the relationship between paternal age and the risk of ASD.

**Fig. 1.**
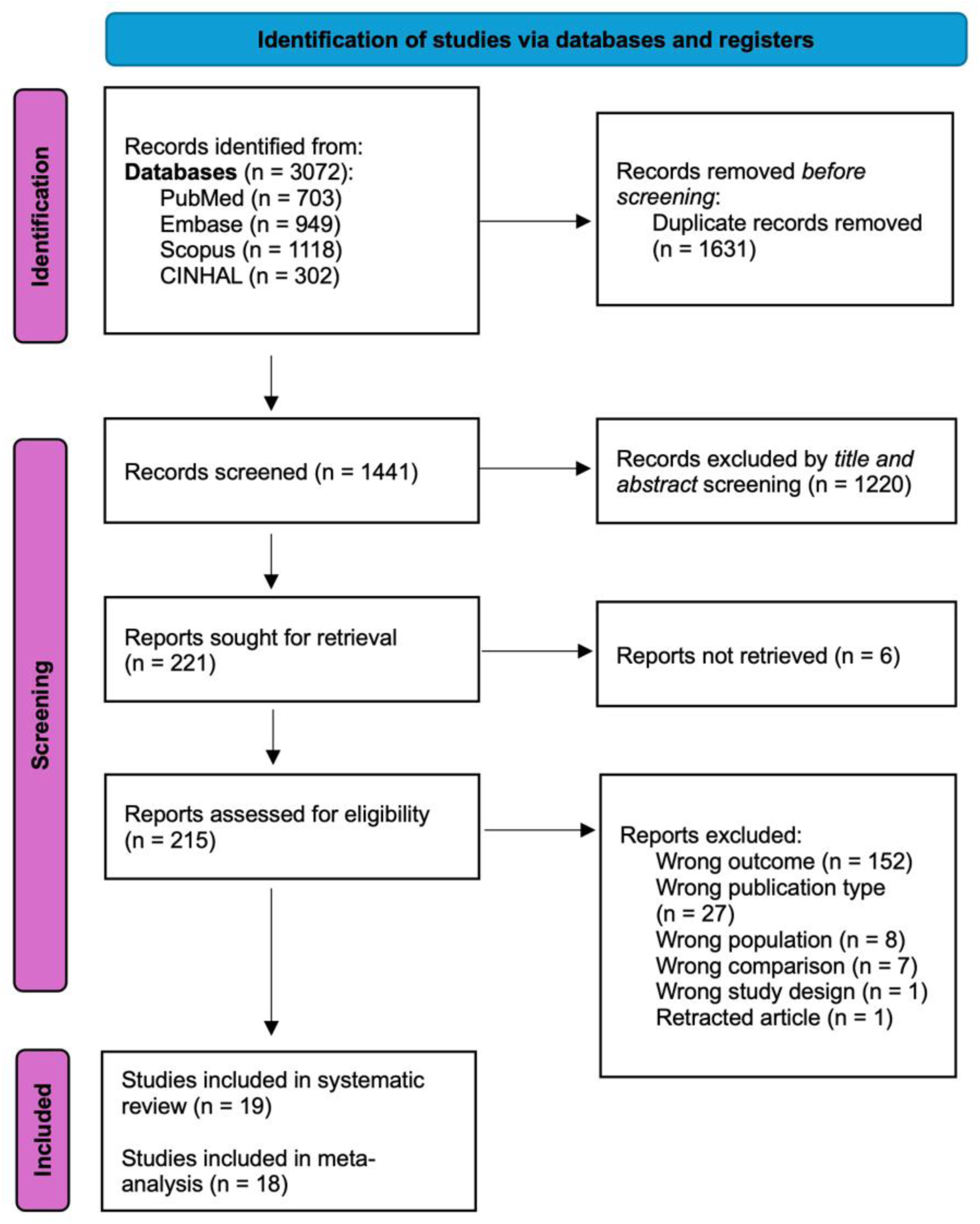
PRISMA flowchart

### 3.2. Characteristics of Included Studies

Table 1 describes all the 18 studies included in the metanalysis from eight MENA countries. The majority of these studies were conducted in Lebanon (n=5 studies), Iran (n=4 studies), Kingdom of Saudi Arabia (KSA) (n=3 studies) and Qatar (n=2 studies). Other countries including Egypt, Jordan, Bahrain, and United Arab Emirates contributed one study each. The included studies were conducted between 2010 and 2024. The total number of participants across all studies was 555,386, with one study (27) contributing most participants (98.9%) to our sample. All 18 studies utilized the case control study design. The Diagnostic and Statistical Manual of Mental Disorders 4^th^ edition (DSM-IV) and 5^th^ edition (DSM-5) were the most used tools (13 studies) for diagnosing ASD in the cases. Other tools used included the Modified Checklist for Autism in Toddlers (M-CHAT) (1 study), Childhood Autism Rating Scale (CARS) (1 study), Autism diagnostic interview (ADI) (3 studies), Social Communication Questionnaire (SCQ) (2 studies), Autism Diagnostic Observation Schedule (ADOS) (1 study), ADOS-2 (1 study) and International Classification of Diseases (ICD) 10^th^ revision (1 study). Finally, in all of the studies except Bener et. al, 2017, over half of the study participants were males.

**Table 1:** Characteristics of Included Studies.

| Study | Year | Country | Type of Study | Number of Participants | Mean Age in Years, ± Standard Deviation (SD) | Assessed Risk Factors | Screening/ Diagnostic Tool | Adjusted Variables | % of Males |
| --- | --- | --- | --- | --- | --- | --- | --- | --- | --- |
| Sasanfar (27) | 2010 | Iran | case-control | 549,533 | 4-11 (Age range) | Consanguinity, male sex, maternal age, paternal age | SCQ, ADI-R, and DSM-IV-TR | Fathers' and mothers' education, birth order, sex, consanguinity, urbanism and province | 51.6% |
| Al-Ansari (28) | 2013 | Bahrain | case-control | 200 | 10.5 ± 6.4 (for cases) | Cesarean Delivery | M-CHAT, DSM-IV-TR and CARS | Age, sex, social class, parents' education, family history | 80% (cases only) |
| Hamade (29) | 2013 | Lebanon | pilot case control | 258 | Cases:<br>12.39 ± 5.92 (boys)<br>10.83 ± 3.23 (girls)<br>Controls:<br>8.09 ± 3.38 (boys)<br>8.45 ± 3.74 (girls) | Male sex | DSM-IV-TR | Living close to an industry, Older age of the child, Male sex of the child, Previous childhood infection, Mother's sadness during pregnancy | 65.1% |
| Bener (30) | 2017 | Qatar | case-control | 616 | 5.39 ± 1.66 (cases)<br>5.62 ± 1.81 (controls) | Consanguinity | ADOS | Age, sex, other variables | 47.1% |
| Guisso (31) | 2018 | Lebanon | case-control | 314 | 8.3 ± 4.8 | Consanguinity, male sex | DSM-IV or DSM-V | Not reported | 64% |
| Oommen (32) | 2018 | KSA | case-control | 200 | 3-10 (Age range) | Consanguinity, maternal age | DSM-V | Age, sex | 64.5% |
| Malek (33) | 2019 | Iran | case-control | 227 | 6.94 ± 3.55 (cases)<br>7.63 ± 3.52 (controls) | Caesarean Delivery, consanguinity, family history, gestational age | DSM-IV-TR and ADI | Age | 60.8% |
| Sadek (34) | 2019 | KSA | case-control | 538 | Not reported | Consanguinity, maternal age, paternal age | DSM-V | Age and sex | 73% |
| Al-Zalabani (35) | 2019 | KSA | case-control | 261 | Not reported | Cesarean Delivery | ICD-10th revision | maternal age at birth, paternal age at birth, maternal education, paternal education, parity, and family income, hypertension, UTI and fever during pregnancy | 74.3% |
| Jenabi (36) | 2020 | Iran | case-control | 300 | 2-10 (Age range) | Cesarean Delivery, male sex, maternal age, gestational age | ADI-R | Not reported | 61.3% |
| Gerges (37) | 2020 | Lebanon | case-control | 200 | 10.14 ± 5.51 (cases)<br>10.40 ± 5.21 (controls) | Family history | DSM-V | Age | 69.5% |
| Shamsedine (38) | 2020 | Lebanon | case-control | 230 | 31.6 months ± 4.9 (cases)<br>25.5 months ± 5.6 (controls) | Family history, paternal age | DSM-V | Sociodemographic, pregnancy, birth variables, parental characteristics, family history | 71.7% |
| Alkhalidy (39) | 2021 | Jordan | case-control | 103 | 4.40 ± 0.80 | Family history | Not reported | All the demographics and familial variables | 61.1% |
| Arafa (40) | 2022 | Egypt | case-control | 772 | 8.82 ± 3.47 (cases)<br>6.42 ± 1.95 (controls) | Cesarean Delivery, Consanguinity, Family history, maternal age | DSM-V | Sex, order of the child, preconception variables, conception variables, and postconception variables | 53% |
| Kheirouri (41) | 2022 | Iran | case-control | 426 | 6.86 ± 1.93 (cases)<br>6.88 ± 1.86 (controls) | Male sex, maternal age, gestational age | DSM-IV-TR | Maternal age, prepregnancy BMI, gestational age, GWG, disease during pregnancy, child's sex, birth weight, birth order, feeding type in first 6 months, maternal and paternal education, and economic status | 66% |
| Hajj (20) | 2022 | Lebanon | case-control | 132 | 2.9±1.57 (Mean age at diagnosis) | Male sex, gestational age | Not reported | Matched for age and mouhafaza | 62.12% |
| Burhani (42) | 2023 | UAE | case-control | 221 | 7.2 ± 3.6 (cases)<br>7.0 ± 2.3 (controls) | Caesarean Delivery, male sex | DSM-V | Obstetric and neonatal risk factors | 76% |
| Alshaban (43) | 2023 | Qatar | case-control | 891 | 8.28 ± 2.70 | Consanguinity | SCQ, ADOS-2, DSM-5 | Paternal age, maternal age, child male sex, child epilepsy | 56.2% |

**Abbreviations.** SCQ, Social Communication Questionnaire; M-CHAT, Modified Checklist for Autism in Toddlers; CARS, Childhood Autism Rating Scale; DSM-IV-TR, Diagnostic and statistical manual of mental disorders, 4th ed, text revision; ADOS, Autism Diagnostic Observation Schedule; ADI, Autism diagnostic interview; *KSA, Kingdom of Saudi Arabia; UAE, United Arab Emirate*

### 3.3. Assessment of the quality of included studies

The included studies were of generally good quality with MASTER scale scores ranging from 21 to 30, and an average quality score of 25 out of 36. The greater the score, the better the quality of a study, indicating that it would have met more safeguards against systematic errors. Most studies included safeguards in five out of the seven areas evaluated by the MASTER scale: equal recruitment, equal retention, implementation, sufficient analysis, and good temporal sequence. However, some studies lacked safeguards in equal ascertainment and equal prognosis. Detailed assessments for each study are available in Supplementary Table S7.

### 3.4. Conception-related Factors and risk of ASD

#### 3.4.1. Maternal Age

Seven studies (26, 27, 32, 34, 36, 40, 41) examined the association between maternal age and ASD. After meta-analysis of 4 of these studies (Fig. 2), the overall effect size suggested no association between maternal age of 35 years or older relative to mothers who were 34 years or younger (OR 1.04, 95%CI 0.71-1.51) with low heterogeneity (I^2^=26.6%, p=0.252). There was little to no evidence of publication bias on the Doi and funnel plots, with an LFK index of 3.10 and Egger’s p-value of 0.277 (Supplementary Fig. S1). Sensitivity analysis suggested that the results remained consistent after successive removal of each study (Supplementary Fig. S2). Three of the 7 studies (26, 32, 41) could not be included in the meta-analysis due to different reasons. One of these (26) studies used a different maternal age cut-off value of 25 years old and found a strong association between maternal age and ASD (OR 13.13, 95%CI 4.63-37.26). The 2 other studies reported the OR for maternal age as a continuous variable. These studies reported opposing association between maternal age and ASD (OR 0.82, 95%CI 0.73-0.91 (41) and OR (1.13, 95%CI 1.05-1.20) (32) (Supplementary Fig. S3).

**Fig. 2.**
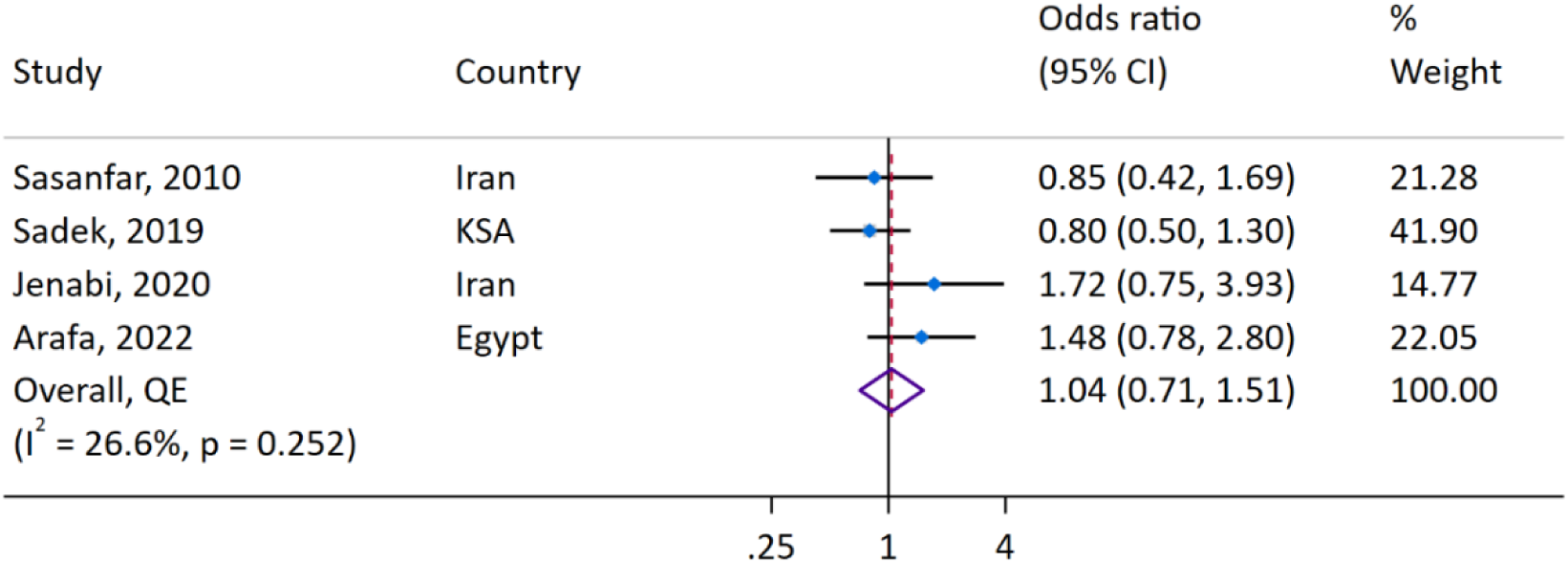
Forest plot of the association between maternal age ≥35 years and ASD

#### 3.4.2. Paternal age

Other risk factors assessed include paternal age and gestational age. However, these studies were not meta-analyzed because of different cut-off values employed by the individual studies in their analyses. Four studies (26, 27, 34, 38) investigated the association between paternal age and ASD. Their results show a trend towards an increase in the risk of ASD with increase in paternal age. Three of the four studies (26, 27, 38) show higher odds of ASD with increasing paternal age, (OR 4.25, 95%CI 1.60-11.28 for paternal age of greater than 35 years, OR 2.03, 95%CI 1.10-3.73 for paternal age of 40 years or more, and OR 1.42, 95%CI 0.64-3.15 for paternal age of 45 years or more). The last study (34) showed no association between paternal age greater than 40 years and ASD (OR 1.00, 95%CI 0.70-1.60). (Supplementary Fig. S4)

### 3.5. Inheritance and Genetic-Related Factors and risk of ASD

#### 3.5.1. Male Sex

As illustrated in Fig. 3, seven studies (20, 27, 29, 31, 36, 41, 42) examined the association between male sex and ASD. Across all seven studies, there was a significant higher odd of ASD in males compared to females. In overall synthesis, male sex, compared to female, was associated with a more than 3-fold increase in the odds of ASD (OR 3.27, 95% CI 2.39-4.48), with low heterogeneity (I^2^=40.9%, p=0.118) (Fig 3). Assessment of publication bias showed no asymmetry on the Doi and funnel plots, with an LFK index of 0.31 and an Egger’s p-value of 0.944 (Supplementary Fig. S5, Supplementary Table S8). Sensitivity analysis showed that results were consistent across all studies (Supplementary Fig. S6).

**Fig. 3.**
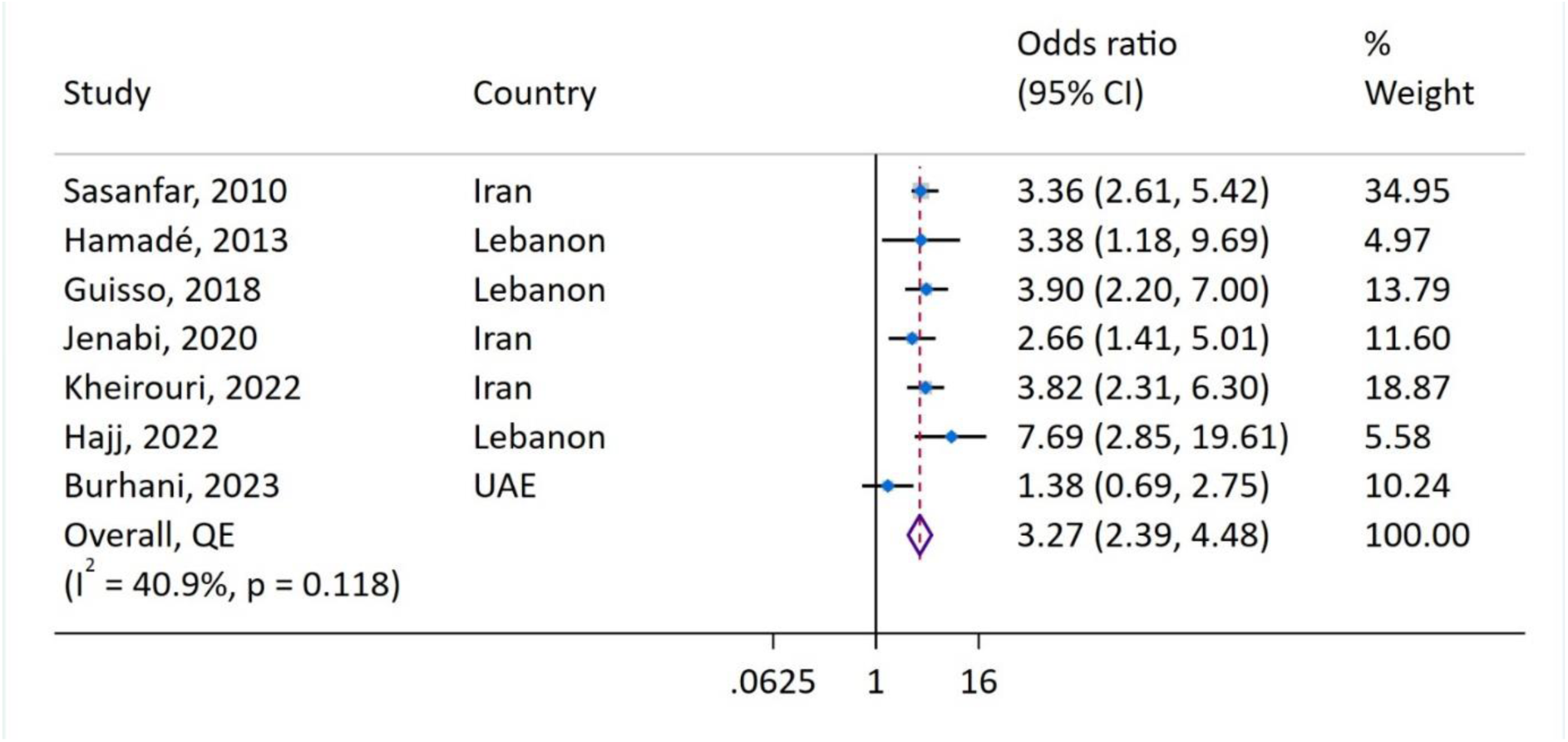
Forest plot of association between male sex and ASD

#### 3.5.2. Consanguinity

Eight studies (27, 30–34, 40, 43) examined the association between consanguinity and ASD (Fig. 4). The overall synthesis of these studies showed an almost 2-fold higher odds of ASD, with consanguinity (OR 1.77, 95% CI 1.38-2.28), with moderate heterogeneity (I^2^=57.1%, p=0.022) and little evidence of publication bias on the Doi and Funnel plots with LFK index=1.64 (Supplementary Fig. S7) and Egger’s p value=0.046 (Supplementary Table S8). Sensitivity analysis showed that results were consistent across all studies (Supplementary Fig. S8).

**Fig. 4.**
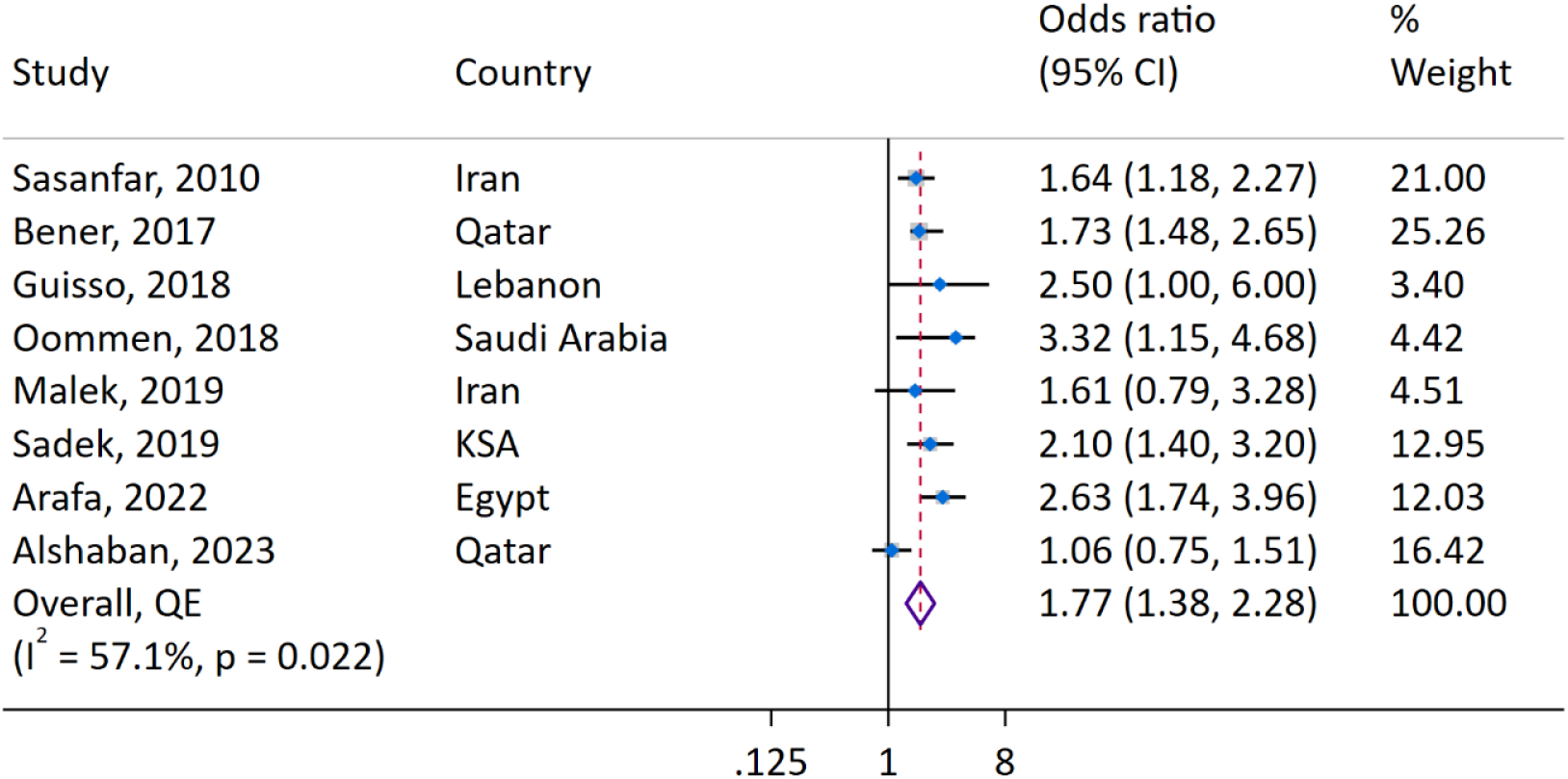
Forest plot showing association between ASD and consanguinity

#### 3.5.3. Family History of ASD

Five studies (33, 37–40) investigated the effect of family history on ASD (Fig. 5). After metanalysis, the overall effect suggested three-fold higher odds of ASD associated with having a family history of ASD (OR 2.98, 95%CI 0.51-17.31) with high heterogeneity (I^2^ =85.0%, p<0.01). Assessment of publication bias showed minor asymmetry on the Doi and funnel plots, with an LFK index of -1.35 (Supplementary Fig. S9) and Egger’s p value of 0.738 (Supplementary Table S8). A leave-one-out analysis showed that one study (39) had the greatest influence on the overall effect size, although the omission of this study did not alter the association observed (Supplementary Fig. S10).

**Fig. 5.**
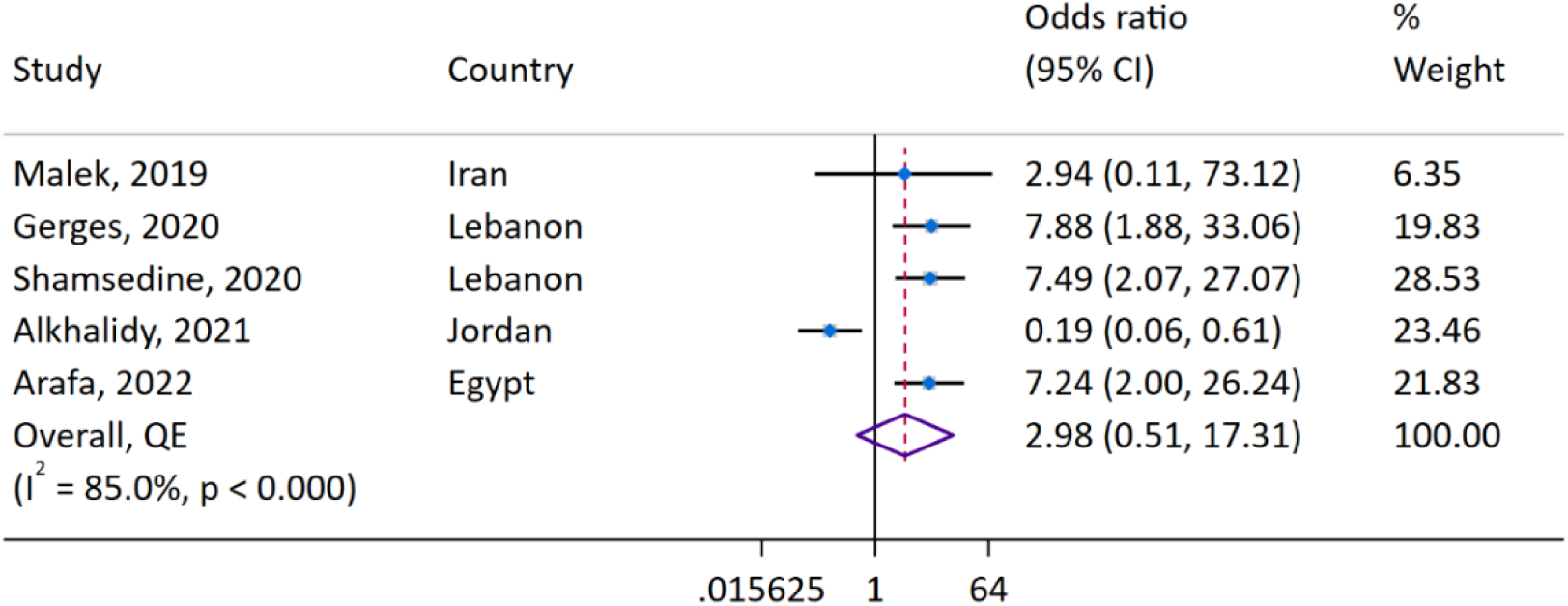
Forest plot of the association between ASD and family history of ASD

### 3.6. Pregnancy Morbidity Factors and risk of ASD

A total of three studies (40–42) investigated the association between pregnancy related comorbidities such as gestational diabetes and maternal hypertension and the risk of ASD. Of these studies, two (40, 41) investigated gestational diabetes, with one (41) reporting an almost 11-fold increase in the odds of ASD (OR 10.76, 95%CI 2.73-42.39) and the other (40), an almost two-fold increase in the odds of ASD (OR 1.75, 95%CI 0.57-5.39) in offspring of women with gestational diabetes. All three studies (40–42) investigated the association of ASD with maternal hypertension, with two (40, 41) reporting a more than three-fold increase in the odds of ASD (OR 3.14, 95%CI 0.93-10.53 and OR 3.64, 95%CI 1.06-12.51 respectively) and the remaining study (42) an almost two-fold increase in the odds of ASD in offspring exposed to maternal hypertension (OR 1.69, 95%CI 0.69-2.75).

### 3.7. Adverse Pregnancy Outcomes and risk of ASD

#### 3.7.1. Cesarean Delivery

Six studies (28, 33, 35, 36, 40, 42) investigated the association between caesarean delivery and ASD, with two studies (28, 35) showing higher odds of ASD for caesarian delivery, one (40) reporting a protective effect of caesarian delivery on ASD and the remaining three studies (33, 36, 42) showing non-significant associations between caesarian delivery and ASD. The meta-analysis of these studies suggested a slightly higher overall odds of ASD with caesarean delivery compared to non-caesarean delivery (OR 1.26, 95%CI 0.67-2.39), with high heterogeneity (I^2^=84.8%, p<0.01) and little evidence of publication bias (Supplementary Fig. S11). A leave-one-out analysis showed that one study (40) had the greatest influence on the overall effect size, although the removal of this study did not alter the conclusions of the analysis (Supplementary Fig. S12).

**Fig. 6.**
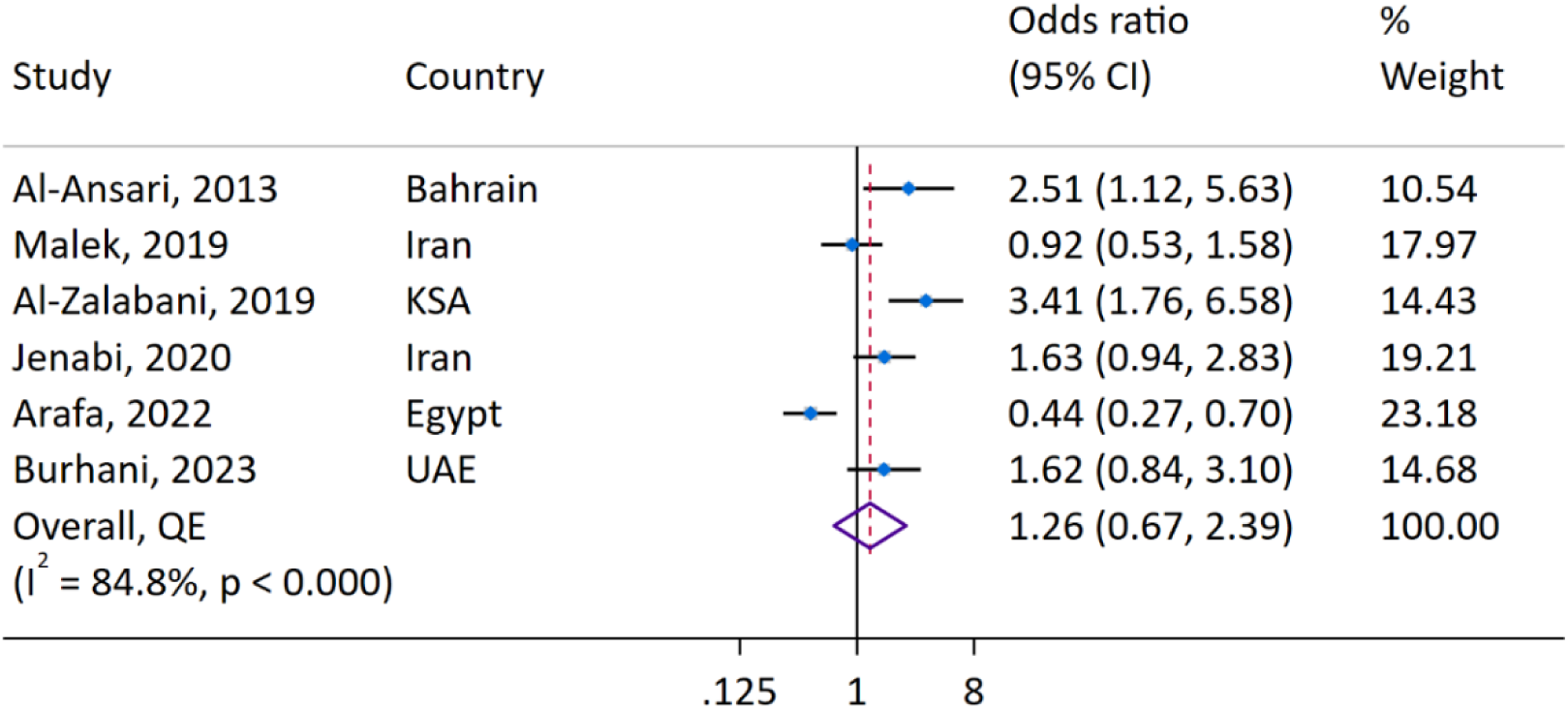
Forest plot showing the association between ASD and cesarean delivery

#### 3.7.2. Gestational age and adverse pregnancy outcomes

Four studies (20, 33, 36, 41) investigated the association between gestational age and ASD (Supplementary Fig. S13). Their results were inconsistent. Two of these studies described the OR for gestational age as a continuous variable and reported opposing ORs of 0.76 (95%CI 0.60-0.96) (20) and 1.21 (95%CI 1.05-1.41) (41) for every unit (one week) increase in gestational age. The two other studies reported the OR for ASD in gestational age of less than 32 weeks and 37 weeks and reported ORs of 0.96 (95%CI 0.43-2.11) (33) and 4.03 (95%CI 1.72-9.42) (36) respectively. Only one study (42) investigated the association between macrosomia and ASD and reported a seven-fold increase in the odds of ASD for children born with macrosomia (OR 6.76, 95%CI 0.76-59.93). For respiratory distress syndrome, only one study (40) investigated this association with ASD, with findings suggesting no association (OR 0.77, 95%CI 0.36-1.66). Lastly, one study (40) investigated the association between neonatal intensive care unit (NICU) admission and ASD, and found a two-fold increase in the odds of ASD for children in NICU (OR 2.13, 95%CI 1.21-3.74)

## 4. Discussion

This systematic review and meta-analysis represents a comprehensive review of 19 published observational studies to date focusing on the risk factors of autism spectrum disorder in the Middle Eastern and North African populations. The results of our meta-analyses suggested that inheritance-related / genetic factors such as male sex, consanguinity, and family history were strongly associated with ASD. A review of the few available studies suggested some associations between ASD and maternal comorbidities during pregnancy as well as some adverse pregnancy complications.

Our review showed some inconsistent results from studies on the association between conception-related factors and the risk of ASD in the MENA region, with some studies showing some associations between older maternal and paternal age and the risk of ASD while others no reported associations. Of these factors, meta-analysis was possible for maternal age ≥35 years only and the overall OR suggested no association with ASD in the MENA region. However, our review of studies with higher cut-offs suggested that older ages may contribute to ASD risk. Two previous meta-analyses, which did not include many studies from the MENA region, have also shown some low-level association between older maternal age and ASD (9, 44). One meta-analysis carried out in 2017 found that maternal age above 35 slightly increased the risk of ASD, with a risk ratio of 1.29 (44). However, substantial heterogeneity was detected in this study with studies from Australia, Sweden, China, and Tunisia reporting no association between this maternal age cut-off and ASD. The other meta-analysis conducted in 2012 reported an almost similar adjusted relative risk for ASD of 1.31 and suggested a dose-response effect of maternal age on the risk of ASD (9). In the current meta-analysis, we were not able to carry out a dose-response analysis due to the sparsity of our data. Advanced maternal age maybe associated with ASD due to chromosomal abnormalities being more likely to be present in the ova with older age (45, 46). In addition, advanced maternal age is also associated with being more likely to have obstetric complications (47). There is the possibility that these complications may increase the risk of ASD, as our review also showed some potential associations between ASD and some maternal comorbidities in pregnancy such as gestational diabetes and gestational hypertension, albeit from a few studies only (48). Therefore, further research is needed, not only on maternal age and risk of ASD but for maternal comorbidities in pregnancy in the region.

The findings from our syntheses showed that, in the MENA region, ASD is strongly associated with inherited / genetic related factors. In overall syntheses, male sex and family history of ASD were both associated with three-fold higher odds of ASD, and consanguinity was associated with a 77% increase in the odds of ASD. The finding for male sex is consistent with findings from two existing global meta-analyses, although these studies reported unadjusted effect sizes and did not include many of our included studies, with one showing a male to female ASD prevalence ratio above four (49) and the other, a risk ratio of 1.47.(44) Although the exact mechanism is unknown, one theory suggests that males have a higher risk of ASD as ASD arises from hypermasculinization of the brain by testosterone (50). It is also important to recognize the possibility of an existing sex-based diagnostic bias, as girls who typically meet the diagnostic criteria for ASD may be less likely to receive a clinical diagnosis, thus exacerbating the observed sex disparity. (49) For consanguinity, we could not find other meta-analyses that investigated this relationship. Consanguinity is known to increase the risk of autosomal recessive conditions and the same relationship could apply to ASD (51). Consanguinity is common in the MENA region, with prevalence ranging from 34% -80% (7) (52) in Saudi Arabia and Qatar and therefore this may be a pertinent risk factor for ASD, unique to the MENA region, warranting inclusion in screening programs. Similar to consanguinity, we could not find existing meta-analyses that examined the association between family history of ASD and ASD. However, data from large cohorts from other regions agree with our finding. For instance, one large population-based cohort study conducted in Sweden that examined 567 436 individuals in Sweden reported nine-fold higher odds of ASD in those with a first-degree relative with ASD (53). Another large population study from Denmark that followed 1,697,231 children reported similar findings (54). Taken together, our findings, and the existing literature suggest that inherited / genetic factors play a strong role in the development of ASD in the MENA region and possibly in other regions too.

Our review suggests that there are possible associations between some adverse pregnancy outcomes such as macrosomia, respiratory distress syndrome, NICU and caesarian section and ASD, although we could only synthesize data for caesarian section. In that synthesis, we found that caesarian delivery increased the odds of ASD by 26% although the 95% confidence interval suggested that there was uncertainty in the estimate of the association. Two previous meta-analyses, which also did not include many studies from the MENA region, found risk estimates almost similar to ours, one showed that cesarean delivery increased the odds of ASD by 33% (55), while the other showed a 25% increase in the odds of ASD (56). Although the reasons are not fully understood, a possible explanation is that this may be due to underlying maternal obstetric complications necessitating a caesarian delivery (57, 58). Another potential explanation suggests that babies born via C-section bypass exposure to the mother’s vaginal bacteria, which could influence gut microbiome development and, in turn, neurodevelopment. In support of this explanation, one study found significant differences exist between the gut microbiome of children with ASD compared to children without ASD (59) Our findings suggest a need for more research on the associations between adverse pregnancy outcomes and the risk of ASD in the MENA region.

Our study has significant strengths. Our meta-analysis included a good number of studies from several countries in the MENA region and may therefore be generalizable across the region. We used a bias-adjusted meta-analysis method that helped us to accommodate poor-quality studies but ensured that their weight contribution to the overall effects were reduced, thus ensuring the use of all available evidence. Nevertheless, the results must still be interpreted with caution as they are based on case-control studies which are subject to potential biases such as recall and selection bias. Additionally, the quality assessment showed that some studies were deficient in some safeguards. Another limitation is that subgroup analysis was not done due to insufficient data and therefore we could not explain the observed heterogeneity where it was noted. Further, we were not able to include some studies in meta-analysis for maternal age and we were not able to carry out meta-analyses for paternal and gestational age due to the variation in how the different studies defined the cut-off points. Finally, the scope of our meta-analysis did not incorporate genetic analyses, which could have been invaluable given ASD’s multifactorial nature. Future research should aim to assess genetic information to carry out a comprehensive analysis of the risk factors associated with ASD.

## 5. Conclusion

These findings confirm the association between male sex, family history of ASD and consanguinity with ASD, potentially suggesting some inherited mechanism in the etiology of ASD in the MENA region. Some maternal co-morbidities during pregnancy and adverse pregnancy outcomes may contribute to ASD risk in the MENA region, although more studies are needed in the region.

## Data Availability

This study is a meta-analysis of published studies and the data used in this work are summary adjusted odds ratios from the included studies. The availability of the underlying individual participant data is subject to the primary publications' journal policies.

## 6. Ethics

This review used secondary data from peer-reviewed published studies and does not require ethical clearance.

## 7. Disclosures

The authors report no conflicts of interest in this work.

## 8. Funding

This work was supported by the Qatar National Research Fund, Undergraduate Research Experience Program (UREP) (Grant ID: UREP30-211-3-073). Open Access Funding provided by the Qatar National Library.

## 9. Author Contributions

Conceptualization: Aishat F. Akomolafe, Fathima R. Mahmood, Tawanda Chivese; Methodology: Aishat F. Akomolafe, Fathima R. Mahmood, Bushra M. Abdallah, Aisha Al-Khulaifi, Yara Dweidri, Duaa Yousif, Nour Darwish, Hafsa Khalid, Majed Al-Theyab, Tawanda Chivese; Writing -Original Draft: Aishat F. Akomolafe, Fathima R. Mahmood, Bushra M. Abdallah, Amgad M. Elshoeibi, Elhassan Mahmoud, Aisha Al-Khulaifi, Yara Dweidri, Duaa Yousif, Nour Darwish, Hafsa Khalid, Majed Al-Theyab; Writing - Review & Editing: Aishat F. Akomolafe, Fathima R. Mahmood, Bushra M. Abdallah, Madeeha Kamal, Majid Alabdulla, Durre Shahwar, Muhammad Waqar Azeem, Salma M. Khaled, Tawanda Chivese; Software: Amgad M. Elshoeibi, Elhassan Mahmoud; Formal analysis: Amgad M. Elshoeibi, Elhassan Mahmoud, Tawanda Chivese; Data Curation: Amgad M. Elshoeibi, Elhassan Mahmoud; Project administration: Aishat F. Akomolafe, Tawanda Chivese, Salma M. Khaled; Funding acquisition: Aishat F. Akomolafe, Tawanda Chivese, Salma M. Khaled; Supervision: Tawanda Chivese, Salma M. Khaled. All authors made a significant contribution to the work reported and gave final approval of the version to be published.

## Notes

### Competing Interest Statement

The authors have declared no competing interest.

