## Supplementary materials for "Developmental origins of the Autism Spectrum Disorder in the Middle East and North Africa Region: A Systematic Review and Meta-Analysis of adjusted risk factors"

[2] Sidra Medicine, Doha, Qatar.

[3] Hamad Medical Corporation, Doha, Qatar.

[4]Weill Cornell Medicine Qatar, Doha, Qatar

Corresponding and guarantor author:

Tawanda Chivese,

Department of Population Medicine, College of Medicine, QU Health, Qatar University, Doha, Qatar

P O BOX 2713

Doha, Qatar

Telephone number: +974 4403 7831

### PRISMA Checklist

Supplementary Table S1. PRISMA Checklist.

| Section and Topic | Item # | Checklist item | Location where item is reported |
|---|---|---|---|
| **TITLE** | | | |
| Title | 1 | Identify the report as a systematic review. | 1 |
| **ABSTRACT** | | | |
| Abstract | 2 | See the PRISMA 2020 for Abstracts checklist. | 2 |
| **INTRODUCTION** | | | |
| Rationale | 3 | Describe the rationale for the review in the context of existing knowledge. | 3 – 4 |
| Objectives | 4 | Provide an explicit statement of the objective(s) or question(s) the review addresses. | 3 – 4 |
| **METHODS** | | | |
| Eligibility criteria | 5 | Specify the inclusion and exclusion criteria for the review and how studies were grouped for the syntheses. | 5 |
| Information sources | 6 | Specify all databases, registers, websites, organisations, reference lists and other sources searched or consulted to identify studies. Specify the date when each source was last searched or consulted. | 4 |
| Search strategy | 7 | Present the full search strategies for all databases, registers and websites, including any filters and limits used. | 4 – 5 |
| Selection process | 8 | Specify the methods used to decide whether a study met the inclusion criteria of the review, including how many reviewers screened each record and each report retrieved, whether they worked independently, and if applicable, details of automation tools used in the process. | 5 |
| Data collection process | 9 | Specify the methods used to collect data from reports, including how many reviewers collected data from each report, whether they worked independently, any processes for obtaining or confirming data from study investigators, and if applicable, details of automation tools used in the process. | 6 |
| Data items | 10a | List and define all outcomes for which data were sought. Specify whether all results that were compatible with each outcome domain in each study were sought (e.g. for all measures, time points, analyses), and if not, the methods used to decide which results to collect. | 5 – 6 |
| | 10b | List and define all other variables for which data were sought (e.g. participant and intervention characteristics, funding sources). Describe any assumptions made about any missing or unclear information. | 5 – 6 |
| Study risk of bias assessment | 11 | Specify the methods used to assess risk of bias in the included studies, including details of the tool(s) used, how many reviewers assessed each study and whether they worked independently, and if applicable, details of automation tools used in the process. | 6 |
| Effect measures | 12 | Specify for each outcome the effect measure(s) (e.g. risk ratio, mean difference) used in the synthesis or presentation of results. | 5-6 |
| Synthesis methods | 13a | Describe the processes used to decide which studies were eligible for each synthesis (e.g. tabulating the study intervention characteristics and comparing against the planned groups for each synthesis (item #5)). | 6 – 7 |
| | 13b | Describe any methods required to prepare the data for presentation or synthesis, such as handling of missing summary statistics, or data conversions. | 6 – 7 |
| | 13c | Describe any methods used to tabulate or visually display results of individual studies and syntheses. | 6 – 7 |
| | 13d | Describe any methods used to synthesize results and provide a rationale for the choice(s). If meta-analysis was performed, describe the model(s), method(s) to identify the presence and extent of statistical heterogeneity, and software package(s) used. | 6 – 7 |

| Section and Topic | Item # | Checklist item | Location where item is reported |
|---|---|---|---|
| | 13e | Describe any methods used to explore possible causes of heterogeneity among study results (e.g. subgroup analysis, meta-regression). | 6 – 7 |
| | 13f | Describe any sensitivity analyses conducted to assess robustness of the synthesized results. | 6 – 7 |
| Reporting bias assessment | 14 | Describe any methods used to assess risk of bias due to missing results in a synthesis (arising from reporting biases). | 6 – 7 |
| Certainty assessment | 15 | Describe any methods used to assess certainty (or confidence) in the body of evidence for an outcome. | N/A |
| **RESULTS** | | | |
| Study selection | 16a | Describe the results of the search and selection process, from the number of records identified in the search to the number of studies included in the review, ideally using a flow diagram. | 7 |
| | 16b | Cite studies that might appear to meet the inclusion criteria, but which were excluded, and explain why they were excluded. | 7 – 8 |
| Study characteristics | 17 | Cite each included study and present its characteristics. | 7 – 11 |
| Risk of bias in studies | 18 | Present assessments of risk of bias for each included study. | 12 |
| Results of individual studies | 19 | For all outcomes, present, for each study: (a) summary statistics for each group (where appropriate) and (b) an effect estimate and its precision (e.g. confidence/credible interval), ideally using structured tables or plots. | 12 – 17 |
| Results of syntheses | 20a | For each synthesis, briefly summarise the characteristics and risk of bias among contributing studies. | 12 – 17 |
| | 20b | Present results of all statistical syntheses conducted. If meta-analysis was done, present for each the summary estimate and its precision (e.g. confidence/credible interval) and measures of statistical heterogeneity. If comparing groups, describe the direction of the effect. | 12 – 17 |
| | 20c | Present results of all investigations of possible causes of heterogeneity among study results. | 12 – 17 |
| | 20d | Present results of all sensitivity analyses conducted to assess the robustness of the synthesized results. | 12 – 17 |
| Reporting biases | 21 | Present assessments of risk of bias due to missing results (arising from reporting biases) for each synthesis assessed. | 12 – 17 |
| Certainty of evidence | 22 | Present assessments of certainty (or confidence) in the body of evidence for each outcome assessed. | N/A |
| **DISCUSSION** | | | |
| Discussion | 23a | Provide a general interpretation of the results in the context of other evidence. | 17-19 |
| | 23b | Discuss any limitations of the evidence included in the review. | 19 |
| | 23c | Discuss any limitations of the review processes used. | 19 |
| | 23d | Discuss implications of the results for practice, policy, and future research. | 18 – 19 |
| **OTHER INFORMATION** | | | |
| Registration and protocol | 24a | Provide registration information for the review, including register name and registration number, or state that the review was not registered. | 4 |
| | 24b | Indicate where the review protocol can be accessed, or state that a protocol was not prepared. | 4 |
| | 24c | Describe and explain any amendments to information provided at registration or in the protocol. | N/A |

| Section and Topic | Item # | Checklist item | Location where item is reported |
|---|---|---|---|
| Support | 25 | Describe sources of financial or non-financial support for the review, and the role of the funders or sponsors in the review. | 20 |
| Competing interests | 26 | Declare any competing interests of review authors. | 20 |
| Availability of data, code and other materials | 27 | Report which of the following are publicly available and where they can be found: template data collection forms; data extracted from included studies; data used for all analyses; analytic code; any other materials used in the review. | 20 |

**Search Strategy**

Supplementary Table S2. PubMed Medline Search. Results: 703. Last updated: 20 January 2024.

| |
|---|
| ("Autism Spectrum Disorder"[Mesh] OR “Autistic Disorder"[Mesh] OR "Asperger Syndrome"[Mesh] OR “autism spectrum disorder” [tiab] OR “ASD” [title] OR “autism spectrum condition” [tiab] OR “ASC” [title] OR “Autism” [tiab] OR “Kanner syndrome” [tiab] OR “autistic disorder” [tiab] OR “childhood autism” [tiab] OR “Asperger syndrome” [tiab] OR “childhood disintegrative disorder” [tiab] OR “pervasive developmental disorder not otherwise specified” [tiab])<br>AND<br>(“MENA” [tiab] OR “Middle East*” [tiab] OR “North Africa*” [tiab] OR “Algeria*” [tiab] OR Bahrain* [tiab] OR Djibouti* [tiab] OR Egypt* [tiab] OR Iran* [tiab] OR Iraq* [tiab] OR Jordan* [tiab] OR Kuwait* [tiab] OR Leban* [tiab] OR Liby* [tiab] OR Malt* [tiab] OR Mauritania* [tiab] OR Morocc* [tiab] OR Oman* [tiab] OR Palestin* [tiab] OR Qatar* [tiab] OR “Saudi” [tiab] OR Somalia* [tiab] OR Sudan* [tiab] OR Syria* [tiab] OR Tunisia* [tiab] OR “United Arab Emirates” [tiab] OR UAE [title] OR Emirati [tiab] OR Yemen* [tiab]) |

Supplementary Table S3. Embase Search. Results: 949. Last updated: 20 January 2024.

| |
|---|
| ('Autism Spectrum Disorder':ti,ab OR 'Autistic Disorder':ti,ab OR 'Asperger Syndrome':ti,ab OR ASD:ti OR 'autism spectrum condition':ti,ab OR ASC:ti OR Autism:ti,ab OR 'Kanner syndrome':ti,ab OR 'childhood autism':ti,ab OR 'childhood disintegrative disorder':ti,ab OR 'pervasive developmental disorder not otherwise specified':ti,ab)<br>AND<br>(MENA:ti,ab OR 'Middle East*':ti,ab OR 'North Africa*':ti,ab OR Algeria*:ti,ab OR Bahrain*:ti,ab OR Djibouti*:ti,ab OR Egypt*:ti,ab OR Iran*:ti,ab OR Iraq*:ti,ab OR Jordan*:ti,ab OR Kuwait*:ti,ab OR Leban*:ti,ab OR Liby*:ti,ab OR Malt*:ti,ab OR Mauritania*:ti,ab OR Morocc*:ti,ab OR Oman*:ti,ab OR Palestin*:ti,ab OR Qatar*:ti,ab OR Saudi:ti,ab OR Somalia*:ti,ab OR Sudan*:ti,ab OR Syria*:ti,ab OR Tunisia*:ti,ab OR 'United Arab Emirates':ti,ab OR UAE:ti OR Emirati:ti,ab OR Yemen*:ti,ab) |

Supplementary Table S4. Scopus Search. Results: 1118. Last updated: 20 January 2024.

| |
|---|
| (TITLE-ABS("autism spectrum disorder") OR TITLE(ASD) OR TITLE-ABS("autism spectrum condition") OR TITLE(ASC) OR TITLE-ABS(Autism) OR TITLE-ABS("Kanner syndrome") OR TITLE-ABS("autistic disorder") OR TITLE-ABS("childhood autism") OR TITLE-ABS("Asperger syndrome") OR TITLE-ABS("childhood disintegrative disorder") OR TITLE-ABS("pervasive developmental disorder not otherwise specified")) |
| AND |
| (TITLE-ABS(MENA) OR TITLE-ABS("Middle East*") OR TITLE-ABS("North Africa*") OR TITLE-ABS(Algeria*) OR TITLE-ABS(Bahrain*) OR TITLE-ABS(Djibouti*) OR TITLE-ABS(Egypt*) OR TITLE-ABS(Iran*) OR TITLE-ABS(Iraq*) OR TITLE-ABS(Jordan*) OR TITLE-ABS(Kuwait*) OR TITLE-ABS(Leban*) OR TITLE-ABS(Liby*) OR TITLE-ABS(Malt*) OR TITLE-ABS(Mauritania*) OR TITLE-ABS(Morocc*) OR TITLE-ABS(Oman*) OR TITLE-ABS(Palestin*) OR TITLE-ABS(Qatar*) OR TITLE-ABS(Saudi) OR TITLE-ABS(Somalia*) OR TITLE-ABS(Sudan*) OR TITLE-ABS(Syria*) OR TITLE-ABS(Tunisia*) OR TITLE-ABS("United Arab Emirates") OR TITLE(UAE) OR TITLE-ABS(Emirati) OR TITLE-ABS(Yemen*)) |

Supplementary Table S5. CINHAL. Results: 302. Last updated: 20 January 2024.

| |
|---|
| ((TI "Autism Spectrum Disorder" OR AB "Autism Spectrum Disorder") OR (TI "Autistic Disorder" OR AB "Autistic Disorder") OR (TI "Asperger Syndrome" OR AB "Asperger Syndrome") OR (TI ASD) OR (TI "autism spectrum condition" OR AB "autism spectrum condition") OR (TI ASC) OR (TI Autism OR AB Autism) OR (TI "Kanner syndrome" OR AB "Kanner syndrome") OR (TI "childhood autism" OR AB "childhood autism") OR (TI "childhood disintegrative disorder" OR AB "childhood disintegrative disorder") OR (TI "pervasive developmental disorder not otherwise specified" OR AB "pervasive developmental disorder not otherwise specified")) |
| AND |
| ((TI MENA OR AB MENA) OR (TI "Middle East*" OR AB "Middle East*") OR (TI "North Africa*" OR AB "North Africa*") OR (TI Algeria* OR AB Algeria*) OR (TI Bahrain* OR AB Bahrain*) OR (TI Djibouti* OR AB Djibouti*) OR (TI Egypt* OR AB Egypt*) OR (TI Iran* OR AB Iran*) OR (TI Iraq* OR AB Iraq*) OR (TI Jordan* OR AB Jordan*) OR (TI |

Kuwait* OR AB Kuwait*) OR (TI Leban* OR AB Leban*) OR (TI Liby* OR AB Liby*) OR (TI Malt* OR AB Malt*) OR (TI Mauritania* OR AB Mauritania*) OR (TI Morocc* OR AB Morocc*) OR (TI Oman* OR AB Oman*) OR (TI Palestin* OR AB Palestin*) OR (TI Qatar* OR AB Qatar*) OR (TI Saudi OR AB Saudi) OR (TI Somalia* OR AB Somalia*) OR (TI Sudan* OR AB Sudan*) OR (TI Syria* OR AB Syria*) OR (TI Tunisia* OR AB Tunisia*) OR (TI "United Arab Emirates" OR AB "United Arab Emirates") OR (TI UAE) OR (TI Emirati OR AB Emirati) OR (TI Yemen* OR AB Yemen*))

**Excluded Reports**

Supplementary Table S6. Excluded reports at full-text screening stage with the reasons for exclusion.

| First Author | Year | Title | Reason for Exclusion |
|---|---|---|---|
| Eapen (1) | 2003 | Epidemiological study of developmental and behavioural disorders in three year old children: A UAE study | Full-text not retrieved |
| Al-Ayadhi (2) | 2005 | Heavy metals and trace elements in hair samples of autistic children in central Saudi Arabia | Wrong outcome |
| Al-Ayadhi (3) | 2005 | Pro-inflammatory cytokines in autistic children in central Saudi Arabia | Wrong outcome |
| Al-Ayadhi (4) | 2005 | Autoimmune connection of autism in Central Saudi Arabia | Wrong outcome |
| Al-Ayadhi (5) | 2005 | Altered oxytocin and vasopressin levels in autistic children in Central Saudi Arabia | Wrong outcome |
| Mostafa (6) | 2008 | Serum anti-myelin - Associated glycoprotein antibodies in Egyptian Autistic children | Wrong outcome |
| Al-Salehi (7) | 2009 | Autism in Saudi Arabia: PresentatioN, Clinical Correlates and Comorbidity | Wrong publication type |
| Walsh (8) | 2010 | High-throughput dna sequencing in autism spectrum disorders (ASD) | Wrong publication type |
| Meguid (9) | 2010 | Reduced serum levels of 25-hydroxy and 1,25-dihydroxy vitamin D in Egyptian children with autism | Wrong outcome |
| El-Ansary (10) | 2010 | Measurement of selected ions related to oxidative stress and energy metabolism in Saudi autistic children | Wrong outcome |
| Al-Farsi (11) | 2011 | Malnutrition among preschool-aged autistic children in Oman | Wrong outcome |
| Al-Yafee (12) | 2011 | Novel metabolic biomarkers related to sulfur-dependent detoxification pathways in autistic patients of Saudi Arabia | Wrong outcome |
| El-Ansary (13) | 2011 | Proinflammatory and proapoptotic markers in relation to mono and di-cations in plasma of autistic patients from Saudi Arabia | Wrong outcome |
| Hussein (14) | 2011 | Characteristics of autism spectrum disorders in a sample of egyptian and saudi patients: Transcultural cross sectional study | Wrong outcome |
| Meguid (15) | 2011 | Evaluation of oxidative stress in autism: Defective antioxidant enzymes and increased lipid peroxidation | Wrong outcome |
| Mohareri (16) | 2011 | Attention deficit hyperactivity symptoms in children with autistic spectrum disorder | Wrong outcome |

| | | | |
|---|---|---|---|
| Abd Elhameed (17) | 2011 | A controlled study of the risk factors and clinical picture of children with Autism in an Egyptian sample | Full-text not retrieved |
| El-Baz (18) | 2011 | Risk factors for autism: An Egyptian study | Wrong outcome |
| Waly (19) | 2011 | Low serum levels of glutathione, homocysteine and total antioxidant capacity are associated with increased risk of autism in Oman | Wrong outcome |
| Amr (20) | 2011 | Sex differences in Arab children with Autism spectrum disorders | Wrong outcome |
| Mohammadi (21) | 2011 | Autism spectrum disorders in Iran | Wrong publication type |
| Masri (22) | 2011 | Profile of developmental delay in children under five years of age in a highly consanguineous community: A hospital-based study - Jordan | Wrong outcome |
| Al-Farsi (23) | 2011 | Brief report: Prevalence of autistic spectrum disorders in the Sultanate of Oman | Wrong outcome |
| Gebril (24) | 2011 | HFE gene polymorphisms and the risk for autism in Egyptian children and impact on the effect of oxidative stress | Wrong outcome |
| El-Ansary (25) | 2011 | Impaired plasma phospholipids and relative amounts of essential polyunsaturated fatty acids in autistic patients from Saudi Arabia | Wrong outcome |
| Zeglam (26) | 2012 | Is there a need for a focused health care service for children with autistic spectrum disorders? A keyhole look at this problem in Tripoli, Libya | Wrong publication type |
| El-Ansary (27) | 2012 | Lipid mediators in plasma of autism spectrum disorders | Wrong outcome |
| El-Ansary (28) | 2012 | Relationship between chronic lead toxicity and plasma neurotransmitters in autistic patients from Saudi Arabia | Wrong outcome |
| Essa (29) | 2012 | Increased markers of oxidative stress in autistic children of the Sultanate of Oman | Wrong outcome |
| Samadi (30) | 2012 | A national study of the prevalence of autism among five-year-old children in Iran | Wrong outcome |
| Zeglam (31) | 2012 | Prevalence of autistic spectrum disorders in Tripoli, Libya: The need for more research and planned services | Wrong outcome |
| Elshahawi (32) | 2012 | Possible association of certain HLA-DRB1 alleles with autism in Egyptian children: Relation to family history of autoimmunity | Wrong publication type |
| Zakareia (33) | 2012 | Study of dual angiogenic/neurogenic growth factors among Saudi autistic children and their correlation with the severity of this disorder | Wrong outcome |
| Amr (34) | 2012 | Comorbid psychiatric disorders in Arab children with Autism spectrum disorders | Wrong outcome |

| | | | |
|---|---|---|---|
| Al-Farsi (35) | 2013 | Impact of nutrition on serum levels of docosahexaenoic acid among Omani children with autism | Wrong outcome |
| Al-Farsi (36) | 2013 | Levels of heavy metals and essential minerals in hair samples of children with autism in Oman: A case-control study | Wrong outcome |
| Al-Rubaye (37) | 2013 | Purine metabolism and oxidative stress in children with autistic spectrum disorders | Wrong outcome |
| Zakareia (38) | 2013 | Evaluation of plasma soluble fatty acid synthase levels among Saudi autistic children: Relation to disease severity | Full-text not retrieved |
| Al-Farsi (39) | 2013 | Association of gestational diabetes mellitus with occurrence of Autism: A cohort study | Wrong publication type |
| Bashir (40) | 2013 | Serum level of desert hedgehog protein in autism spectrum disorder: Preliminary results | Wrong outcome |
| Hashim (41) | 2013 | Association between plasma levels of transforming growth factor-β1, IL-23 and IL-17 and the severity of autism in Egyptian children | Wrong outcome |
| Mousavizadeh (42) | 2013 | Association of human mtDNA mutations with autism in Iranian patients | Wrong publication type |
| Kaddah (43) | 2013 | Screening for autism in low-birth-weight Egyptian toddlers | Wrong population |
| Al-Hakbany (44) | 2014 | The Relationship of HLA Class I and II Alleles and Haplotypes with Autism: A Case Control Study | Wrong outcome |
| Ranjbar (45) | 2014 | Comparison of urinary oxidative biomarkers in Iranian children with autism | Wrong outcome |
| Hodgson (46) | 2014 | Decreased glutathione and elevated hair mercury levels are associated with nutritional deficiency-based autism in Oman | Wrong outcome |
| Yassa (47) | 2014 | Autism: A form of lead and mercury toxicity | Wrong outcome |
| Alabdali (48) | 2014 | Association of social and cognitive impairment and biomarkers in autism spectrum disorders | Wrong outcome |
| El-Ansary (49) | 2014 | Role of amino acids in the pathophysiology of autism spectrum disorders in Saudi and Egyptian population samples | Wrong outcome |
| Shawky (50) | 2014 | Study of genotype-phenotype correlation of methylene tetrahydrofolate reductase (MTHFR) gene polymorphisms in a sample of Egyptian autistic children | Wrong outcome |
| Afsharpaiman (51) | 2014 | An assessment of toxoplasmosis antibodies seropositivity in children suffering autism | Wrong outcome |
| Mohamed (52) | 2015 | Assessment of Hair Aluminum, Lead, and Mercury in a Sample of Autistic Egyptian Children: Environmental Risk Factors of Heavy Metals in Autism | Wrong outcome |

| | | | |
|---|---|---|---|
| Halepoto (53) | 2015 | Correlation Between Hedgehog (Hh) Protein Family and Brain-Derived Neurotrophic Factor (BDNF) in Autism Spectrum Disorder (ASD) | Wrong outcome |
| Fanid (54) | 2015 | Association between common single- nucleotide polymorphism of reelin gene, rs736707 (C/T) with autism spectrum disorder in Iranian-Azeri patients | Wrong outcome |
| Fernell (55) | 2015 | Autism spectrum disorder and low vitamin D at birth: A sibling control study | Wrong population |
| Slama (56) | 2015 | Family history of psychiatric disorder and autism spectrum disorders: A study about 790 cases | Wrong publication type |
| Meguid (57) | 2015 | Evaluation of MTHFR genetic polymorphism as a risk factor in Egyptian autistic children and mothers | Wrong outcome |
| Mousavi (58) | 2015 | RoRa gene contribution to autism; another epigenetic layer on autism complexity | Wrong outcome |
| Ouhtit (59) | 2015 | Underlying factors behind the low prevalence of autism spectrum disorders in oman sociocultural perspective | Wrong publication type |
| Yavarna (60) | 2015 | High diagnostic yield of clinical exome sequencing in Middle Eastern patients with Mendelian disorders | Wrong outcome |
| Khakzad (61) | 2015 | Transforming growth factor beta 1 869T/C and 915G/C polymorphisms and risk of autism spectrum disorders | Wrong outcome |
| Afrazeh (62) | 2015 | Measurement of Serum Superoxide Dismutase and Its Relevance to Disease Intensity Autistic Children | Wrong outcome |
| Saad (63) | 2015 | ADHD, autism and neuroradiological complications among phenylketonuric children in Upper Egypt | Wrong outcome |
| Al-Sharbati (64) | 2016 | Profile of mental and behavioral disorders among preschoolers in a tertiary care hospital in Oman: A retrospective study | Wrong outcome |
| Fahmy (65) | 2016 | Vitamin D intake and sun exposure in autistic children | Wrong outcome |
| Chaaya (66) | 2016 | Prevalence of Autism Spectrum Disorder in Nurseries in Lebanon: A Cross Sectional Study | Wrong outcome |
| Haghiri (67) | 2016 | Analysis of methionine synthase (rs1805087) gene polymorphism in autism patients in northern Iran | Wrong outcome |
| El-Ansary (68) | 2016 | Data of multiple regressions analysis between selected biomarkers related to glutamate excitotoxicity and oxidative stress in Saudi autistic patients | Wrong outcome |
| No author name (69) | 2016 | Environmental toxic pollutant and trace elements in Egyptian children with autism | Full-text not retrieved |

| | | | |
|---|---|---|---|
| Dinkler (70) | 2016 | Maltreatment-associated neurodevelopmental problems: Environment and genetics | Wrong population |
| Elhawary (71) | 2016 | Vulnerability of genetic variants to the risk of autism among Saudi children | Wrong outcome |
| Fanid (72) | 2016 | An Association Analysis of Reelin Gene (RELN) exon 22 (G/C), Rs.362691, polymorphism with autism spectrum disorder among Iranian-Azeri population | Wrong outcome |
| Khaled (73) | 2016 | Altered urinary porphyrins and mercury exposure as biomarkers for autism severity in Egyptian children with autism spectrum disorder | Wrong outcome |
| Meguid (74) | 2016 | Impact of oxidative stress on autism spectrum disorder behaviors in children with autism | Full-text not retrieved |
| Mohammed (75) | 2016 | Incidence of autism in high risk neonatal follow up | Wrong population |
| Alhowikan (76) | 2017 | Secretagogin (SCGN) plasma levels and their association with cognitive and social behavior in children with autism spectrum disorder (ASD) | Wrong outcome |
| Alshaban (77) | 2017 | Autism spectrum disorder in Qatar: Profiles and correlates of a large clinical sample | Wrong outcome |
| Desoky (78) | 2017 | Biochemical assessments of thyroid profile, serum 25-hydroxycholecalciferol and cluster of differentiation 5 expression levels among children with autism | Wrong outcome |
| Ashaat (79) | 2017 | Altered adaptive cellular immune function in a group of Egyptian children with autism | Wrong outcome |
| Firouzabadi (80) | 2017 | Copy Number Variants in Patients with Autism and Additional Clinical Features: Report of VIPR2 Duplication and a Novel Microduplication Syndrome | Wrong outcome |
| Khaniani (81) | 2017 | Autistic Phenotype of Permutation and Intermediate Alleles of FMR1 Gene | Wrong outcome |
| Kourtian (82) | 2017 | Candidate Genes for Inherited Autism Susceptibility in the Lebanese Population | Wrong outcome |
| Zeglam (83) | 2017 | Epidemiology of autism in Libya: Uncovering of the first autism findings | Wrong publication type |
| Dinkler (84) | 2017 | Maltreatment-associated neurodevelopmental disorders: a co-twin control analysis | Wrong population |
| Ajabi (85) | 2017 | A study of MTRR 66A>G gene polymorphism in patients with autism from northern Iran | Wrong outcome |
| Hosseinpour (86) | 2017 | Neuropilin-2 rs849563 gene variations and susceptibility to autism in Iranian population: A case-control study | Wrong outcome |
| Hamedani (87) | 2017 | Ras-like without CAAX 2 (RIT2): a susceptibility gene for autism spectrum disorder | Wrong outcome |
| El-Ansary (88) | 2017 | Relationship between selenium, lead, and mercury in red blood cells of Saudi autistic children | Wrong outcome |

| | | | |
|---|---|---|---|
| Noroozi (89) | 2017 | Association study of the vesicular monoamine transporter 1 (VMAT1) gene with autism in an Iranian population | Wrong outcome |
| Safari (90) | 2017 | Synaptosome-Associated Protein 25 (SNAP25) Gene Association Analysis Revealed Risk Variants for ASD, in Iranian Population | Wrong outcome |
| Sayad (91) | 2017 | Retinoic acid-related orphan receptor alpha (RORA) variants are associated with autism spectrum disorder | Wrong outcome |
| Zare (92) | 2017 | The association of CNTNAP2 rs7794745 gene polymorphism and autism in Iranian population | Wrong outcome |
| Zeglam (93) | 2017 | Early TV viewing and autistic spectrum disorder; A plausible hypothesis that should not be dismissed "Libyan Viewpoint" | Wrong publication type |
| Abdulamir (94) | 2018 | Acetylserotonin O-Methyltransferase (ASMT)/rs4446909 Polymorphism in Iraqi Autistic Children | Wrong outcome |
| Kadhim (95) | 2018 | Sero-positivity rate of rubella antibodies in Iraqi autistic children | Wrong outcome |
| Belkady (96) | 2018 | Chromosomal Abnormalities in Patients with Intellectual Disability: A 21-Year Retrospective Study | Wrong outcome |
| Delgado (97) | 2018 | Role of Metal Ion Dyshomeostasis in ASD: Evaluation of Copper, Zinc, and Selenium Levels in the North American ASD Population | Wrong publication type |
| Alshiban (98) | 2018 | Risk factors for Autism Spectrum Disorder (ASD) in Saudi Arabia | Wrong publication type |
| Mousavi (99) | 2018 | Autism and probable prerequisites: Severe and scheduled prenatal stresses at spotlight | Wrong outcome |
| Yousefian (100) | 2018 | Long-term exposure to ambient air pollution and autism spectrum disorder in children: A case-control study in Tehran, Iran | Wrong outcome |
| John (101) | 2018 | Is the prevalence of autism spectrum disorder decreased in black and ethnic children and adolescents? | Wrong publication type |
| Abdulamir (102) | 2018 | Serotonin and serotonin transporter levels in autistic children | Wrong outcome |
| Meguid (103) | 2018 | Frequency of risk factors and coexisting abnormalities in a population of Egyptian children with autism spectrum disorder | Wrong outcome |
| Olusanya (104) | 2018 | Developmental disabilities among children younger than 5 years in 195 countries and territories, 1990–2016: a systematic analysis for the Global Burden of Disease Study 2016 | Wrong population |
| Altamimi (105) | 2018 | Could Autism Be Associated With Nutritional Status in the Palestinian population? The Outcomes of the Palestinian Micronutrient Survey | Wrong study design |

| | | | |
|---|---|---|---|
| Noroozi (106) | 2018 | Association analysis of the GABRB3 promoter variant and susceptibility to autism spectrum disorder | Wrong outcome |
| Qasem (107) | 2018 | Impaired lipid metabolism markers to assess the risk of neuroinflammation in autism spectrum disorder | Wrong outcome |
| Arastoo (108) | 2018 | Evaluation of serum 25-Hydroxy vitamin D levels in children with autism spectrum disorder | Wrong outcome |
| Jabbar (109) | 2018 | Study of polymorphism in methionine synthase gene by RFLP-PCR in middle euphrates region of Iraq | Wrong outcome |
| Eftekharian (110) | 2018 | Expression Analysis of Protein Inhibitor of Activated STAT (PIAS) Genes in Autistic Patients | Wrong outcome |
| Sayad (111) | 2018 | Association of HLA alleles with autism | Wrong outcome |
| Al-Mamri (112) | 2019 | Revisiting the prevalence of autism spectrum disorder among omani children a multicentre study | Wrong outcome |
| Alawad (113) | 2019 | Lead among children with autism in Iraq. Is it a potential factor?: Lead level in Iraqi children with autism | Wrong outcome |
| Alshaban (114) | 2019 | Prevalence and correlates of autism spectrum disorder in Qatar: a national study | Wrong outcome |
| Alzghoul (115) | 2019 | The association between levels of inflammatory markers in autistic children compared to their unaffected siblings and unrelated healthy controls | Wrong outcome |
| Elsayed (116) | 2019 | Study of autistic features among children and adolescents with congenital adrenal hyperplasia | Wrong population |
| Jabbar (117) | 2019 | Evaluation the Relationship Between DRD1 RS4532 Gene Polymorphisms and Autism by RFLP-PCR in Middle Euphrates Region of Iraq | Wrong outcome |
| Manzouri (118) | 2019 | Advanced parental age and risk of positive autism spectrum disorders screening | Wrong outcome |
| Alhader (119) | 2019 | The potential interactive role of leptin with other hormones in the pathophysiology of asd in jordanian male children | Wrong publication type |
| Arab (120) | 2019 | Methylenetetrahydrofolate reductase gene variants confer potential vulnerability to autism spectrum disorder in a Saudi community | Wrong outcome |
| El Khatib (121) | 2019 | Gastrointestinal symptoms in children with autism spectrum disorders and correlation with autism functionality and severity: a case-control study | Wrong publication type |
| Ismail (122) | 2019 | Study of C677T variant of methylene tetrahydrofolate reductase gene in autistic spectrum disorder Egyptian children | Wrong outcome |

| | | | |
|---|---|---|---|
| Mohammadi (123) | 2019 | Prevalence of Autism and its Comorbidities and the Relationship with Maternal Psychopathology: A National Population-Based Study | Wrong outcome |
| Sayad (124) | 2019 | Association study of sequence variants in voltage-gated Ca2+ channel subunit alpha-1C and autism spectrum disorders | Wrong outcome |
| Bordeleau (125) | 2019 | Microglia as a potential link between pathological myelination and stereotypic behavior after exposure to maternal high-fat diet | Wrong publication type |
| Goodarzi (126) | 2019 | Evaluation of autistic spectrum disorders screening in children of Khorramabad (West of Iran) between 2015 and 2016 | Wrong outcome |
| Gleeson (127) | 2019 | The genetic and molecular basis of neurodevelopmental disorders | Wrong publication type |
| Hasan (128) | 2019 | Assessment of children upon suffering from withdrawals signs in Baghdad city, Iraq | Wrong outcome |
| Alzghoul (129) | 2020 | The Association Between Serum Vitamin D3 Levels and Autism Among Jordanian Boys | Wrong outcome |
| Hussien (130) | 2020 | Evaluation of lead, copper and zinc levels for autistic children in thi-qar Province/Iraq | Wrong outcome |
| Khalil (131) | 2020 | Assessing risk factors of Autism Spectrum Disorders (ASD) and Attention Deficit Hyperactivity Disorder (ADHD) among Saudi Mothers: A retrospective study | Wrong publication type |
| Mossa (132) | 2020 | Evaluation of serum oxytocin hormone level in children with autism | Wrong outcome |
| Razjouyan (133) | 2020 | A Study of the Prevalence of Risk Factors Associated with Autism Spectrum Disorder among Affected Individuals in Tehran | Wrong comparison |
| Richa (134) | 2020 | Estimating the prevalence of autism spectrum disorder in Lebanon | Wrong outcome |
| Virolainen (135) | 2020 | Autism spectrum disorder in the United Arab Emirates: Potential environmental links | Wrong publication type |
| Muftin (136) | 2020 | Identification of MTHFR genetic polymorphism in Iraqi autistic children | Wrong outcome |
| Beiranvandi (137) | 2020 | The association of CNTNAP2 rs2710102 and ENGRAILED-2 rs1861972 genes polymorphism and autism in Iranian population | Wrong outcome |
| Darvish (138) | 2020 | Association of rs3735025 and rs9656169 variants with autism, and schizophrenia: A GWAS-replication study in an Iranian population | Wrong outcome |
| Kandeel (139) | 2020 | Impact of Clostridium Bacteria in Children with Autism Spectrum Disorder and Their Anthropometric Measurements | Wrong outcome |
| Mobasheri (140) | 2020 | Association between vitamin D receptor gene FokI and TaqI variants with autism spectrum disorder predisposition in Iranian population | Wrong outcome |

| | | | |
|---|---|---|---|
| Saad (141) | 2020 | Polymorphism of interleukin-1β and interleukin-1 receptor antagonist genes in children with autism spectrum disorders | Wrong outcome |
| Safari (142) | 2020 | The rs12826786 in HOTAIR lncRNA Is Associated with Risk of Autism Spectrum Disorder | Wrong outcome |
| Taheri (143) | 2020 | The rs594445 in MOCOS gene is associated with risk of autism spectrum disorder | Wrong outcome |
| Al-Bazzaz (144) | 2020 | Estimation of fasting serum levels of glucose, zinc, copper, zinc /copper ratio and their relation to the measured lipid profile in autistic patients and non-autistic controls in Jordan | Wrong outcome |
| Chehbani (145) | 2020 | The status of chemical elements in the blood plasma of children with autism spectrum disorder in Tunisia: a case-control study | Wrong outcome |
| Sabbagh (146) | 2021 | Prevalence and characteristics of autistic children attending autism centres in 2 major cities in saudi arabia | Wrong outcome |
| Alomar (147) | 2021 | Vitamin D deficient diet and autism | Wrong outcome |
| Al-Ali (148) | 2021 | Determination of environmental risk factors of Autism in Kerbala city / Iraq 2020 | Wrong outcome |
| Al-Sarraj (149) | 2021 | Family-based genome-wide association study of autism spectrum disorder in middle eastern families | Wrong outcome |
| Hegazy (150) | 2021 | Environmental risk factors associated with children autism spectrum disorders in, menoufia governorate | Wrong outcome |
| Mondal (151) | 2021 | Role of glucose 6-phosphate dehydrogenase (G6PD) deficiency and its association to Autism Spectrum Disorders | Wrong outcome |
| Nawaz (152) | 2021 | Low Birth Weight Prevalence in Children Diagnosed with Neurodevelopmental Disorders in Dubai | Wrong outcome |
| Yousef (153) | 2021 | Prevalence and risk factors of autism spectrum disorders in preschool children in Sharkia, Egypt: a community-based study | Wrong outcome |
| Al Malki (154) | 2021 | Maternal toxoplasmosis and the risk of childhood autism: serological and molecular small-scale studies | Wrong outcome |
| Meguid (155) | 2021 | Awareness and risk factors of autism spectrum disorder in an Egyptian population | Wrong comparison |
| Rahmani (156) | 2021 | Genetic and molecular biology of autism spectrum disorder among Middle East population: a review | Wrong publication type |
| Nakhla (157) | 2021 | Assessment of 25 Hydroxy Cholecalceferol Level in Autistic Children; is there a role for it in Treatment of Autism Spectrum Disorder? | Wrong outcome |

| | | | |
|---|---|---|---|
| Shehata (158) | 2021 | Comparing levels of urinary phthalate metabolites in egyptian children with autism spectrum disorders and healthy control children: Referring to sources of phthalate exposure | Wrong outcome |
| Zaky (159) | 2021 | Neurodevelopmental Outcomes after Neonatal Mechanical Ventilation | Wrong publication type |
| Alotaibi (160) | 2021 | Sociodemographic, clinical characteristics, and service utilization of young children diagnosed with autism spectrum disorder at a research center in Saudi Arabia | Wrong comparison |
| Sadek (161) | 2021 | Clinical and laboratory characteristics of children with autism spectrum disorder at sohag university hospital | Wrong outcome |
| Alrahili (162) | 2021 | The Association Between Screen Time Exposure and Autism Spectrum Disorder-Like Symptoms in Children | Wrong outcome |
| Zahra (163) | 2022 | Socio-economic Status in Egyptian Patients with Autism Spectrum Disorder. Does it affect Autism Severity? | Wrong comparison |
| Aloufi (164) | 2022 | Breastfeeding and Its Relation with Autism Spectrum Disorder in Children | Wrong outcome |
| AlBatti (165) | 2022 | Prevalence of autism spectrum disorder among Saudi children between 2 and 4 years old in Riyadh | Wrong outcome |
| Gholamalizadeh (166) | 2022 | The association of body mass index and dietary fat intake with autism in children: a case-control study | Retracted article |
| Sadiq (167) | 2022 | Toxoplasmosis is a risk factor in autism disease in Al-Diwaniyah governorate, Iraq | Wrong outcome |
| Slama (168) | 2022 | Risk factors in autism spectrum disorder: A Tunisian case-control study | Wrong outcome |
| Akbari (169) | 2022 | Association between angiotensin I converting enzyme gene polymorphisms and risk of autism in Iranian population | Wrong outcome |
| Lord (170) | 2022 | How matrix metalloproteinase (MMP)-9 (rs3918242) polymorphism affects MMP-9 serum concentration and associates with autism spectrum disorders: A case-control study in Iranian population | Wrong outcome |
| Hamed (171) | 2022 | Anti-ganglioside M1 autoantibodies in Egyptian children with autism: a cross-sectional comparative study | Wrong outcome |
| Hassan (172) | 2022 | Vitamin D3 status and polymorphisms of vitamin D receptor genes among cohort of Egyptian children with autism | Wrong outcome |
| Raouf (173) | 2022 | Association of immune abnormalities with symptom severity in Egyptian autistic children | Wrong outcome |

| | | | |
|---|---|---|---|
| Rezaei (174) | 2022 | A case-control study on the relationship between urine trace element levels and autism spectrum disorder among Iranian children | Wrong outcome |
| Al-Ali (175) | 2022 | The oxytocin receptor gene polymorphism rs2268491 and serum oxytocin alterations are indicative of autism spectrum disorder: A case-control paediatric study in Iraq with personalized medicine implications | Wrong outcome |
| Al-Dakroury (176) | 2022 | Autism in the Kingdom of Saudi Arabia: Current Situation and Future Perspectives for Services and Research | Wrong publication type |
| Aljumaili (177) | 2022 | Assessment of hair aluminium, cobalt, and mercury in a specimen of autistic Iraqi patients: Environmental risk factors of heavy metals in autism | Wrong outcome |
| Alenezi (178) | 2022 | Psychotropic Medications Use among Children with Autism in Saudi Arabia | Wrong comparison |
| Dehiol (179) | 2022 | Autism spectrum disorders and electronic screen devices exposure in Al-Nasiriya city 2019-2020 | Wrong outcome |
| Aldera (180) | 2022 | Do Parental Comorbidities Affect the Severity of Autism Spectrum Disorder? | Wrong outcome |
| Sefrioui (181) | 2023 | Profile of autism spectrum disorders in Morocco: cross-sectional retrospective study of parents of children with autism | Wrong outcome |
| Alakhzami (182) | 2023 | Individuals with Autism Spectrum Disorders and Developmental Disorders in Oman: An Overview of Current Status | Wrong publication type |
| Alamoudi (183) | 2023 | Prenatal maternal stress and the severity of autism spectrum disorder: A cross-sectional study | Wrong comparison |
| Al-Awadi (184) | 2023 | Chromosomal aberration detection in Iraqi children with autism | Wrong outcome |
| Aljumaili (185) | 2023 | Determination of hair lead, iron, and cadmium in a sample of autistic Iraqi children: Environmental risk factors of heavy metals in autism | Wrong outcome |
| AlQahtani (186) | 2023 | Autism spectrum disorder: Where does the Gulf Region stand? An overview of ASD in the Arab Gulf Region: The UAE as a regional model | Wrong publication type |
| Heidari (187) | 2023 | The Association Between Autism Spectrum Disorder and Attention Deficit Hyperactivity Disorder Symptoms in Medical Students | Wrong outcome |
| Jenabi (188) | 2023 | Autism Spectrum Disorders Registry in Hamadan, Iran: A Study Protocol | Wrong publication type |
| Jenabi (189) | 2023 | Is Breastfeeding Duration Associated with Risk of Developing ASD? | Full-text not retrieved |
| Kassab (190) | 2023 | Assessment of caries prevalence and related risk factors among a group of lebanese children with autism spectrum disorder: a case-control study | Wrong outcome |

| | | | |
|---|---|---|---|
| Meguid (191) | 2023 | Prevalence of autism spectrum disorder among children referred to special needs clinic in Giza | Wrong population |
| Nasir (192) | 2023 | Pediatricians' perspectives on childhood behavioral and mental health problems in Jordan | Wrong outcome |
| Saleh (193) | 2023 | Hair and Blood Levels of Aluminum, Cadmium, and Lead in Children with Autism from Egypt: Can Toxic Heavy Metals Increase the Risk of Autism? | Wrong outcome |
| Al-Bishri (194) | 2023 | Glucose transporter 1 deficiency, AMP-activated protein kinase activation and immune dysregulation in autism spectrum disorder: Novel biomarker sources for clinical diagnosis | Wrong outcome |
| Hassan (195) | 2023 | Toxoplasmosis and cytomegalovirus infection and their role in Egyptian autistic children | Wrong outcome |
| Sadeghi (196) | 2023 | Associations between Symptom Severity of Autism Spectrum Disorder and Screen Time among Toddlers Aged 16 to 36 Months | Wrong outcome |
| Meimand (197) | 2023 | Burden of autism spectrum disorders in North Africa and Middle East from 1990 to 2019: A systematic analysis for the Global Burden of Disease Study 2019 | Wrong outcome |
| Abdi (198) | 2023 | Genomic architecture of autism spectrum disorder in Qatar: The BARAKA-Qatar Study | Wrong outcome |
| Moravej (199) | 2023 | Inborn Errors of Metabolism Associated With Autism Among Children: A Multicenter Study from Iran | Wrong comparison |
| Boujelben (200) | 2023 | Familial Autism Spectrum Disorder : A clinical study from South Tunisia | Wrong publication type |
| Ghahari (201) | 2023 | Prenatal exposure to ambient air pollution and autism spectrum disorders: Results from a family-based case-control study | Wrong outcome |
| Zebbiche (202) | 2024 | Trace element levels and autism spectrum disorder in a sample of Algerian children: A case-control study investigation | Wrong outcome |

### Assessment of the Quality of Included Studies

Supplementary Table S7. Scores for the assessment of the quality of included studies using the MASTER Scale.

| Study | Equal recruitment | | | | Equal retention | | | | | Equal ascertainment | | | | | | | Equal implementation | | | | | | Equal prognosis | | | | | | Sufficient analysis | | | Temporal precedence | | | | | Total |
|---|---|---|---|---|---|---|---|---|---|---|---|---|---|---|---|---|---|---|---|---|---|---|---|---|---|---|---|---|---|---|---|---|---|---|---|---|---|
| | 1 | 2 | 3 | 4 | 5 | 6 | 7 | 8 | 9 | 10 | 11 | 12 | 13 | 14 | 15 | 16 | 17 | 18 | 19 | 20 | 21 | 22 | 23 | 24 | 25 | 26 | 27 | 28 | 29 | 30 | 31 | 32 | 33 | 34 | 35 | 36 | |
| Sasanfar, 2010 | 1 | 0 | 1 | 1 | 1 | 1 | 1 | 1 | 1 | 1 | 1 | 1 | 0 | 0 | 0 | 0 | 1 | 1 | 0 | 1 | 1 | 1 | 1 | 1 | 0 | 0 | 0 | 1 | 1 | 1 | 1 | 0 | 1 | 1 | 1 | 1 | 26 |
| Al-Ansari, 2013 | 1 | 1 | 1 | 1 | 1 | 1 | 1 | 1 | 1 | 1 | 1 | 1 | 0 | 0 | 0 | 0 | 0 | 1 | 0 | 1 | 1 | 1 | 1 | 1 | 1 | 0 | 0 | 1 | 1 | 1 | 1 | 0 | 1 | 1 | 1 | 1 | 27 |
| Hamadé, 2013 | 1 | 0 | 1 | 1 | 1 | 1 | 1 | 1 | 1 | 1 | 1 | 1 | 1 | 0 | 0 | 0 | 0 | 1 | 0 | 1 | 1 | 1 | 1 | 1 | 1 | 0 | 0 | 1 | 1 | 1 | 1 | 0 | 1 | 1 | 1 | 1 | 27 |
| Bener, 2017 | 1 | 1 | 1 | 1 | 0 | 1 | 1 | 1 | 1 | 1 | 1 | 0 | 0 | 0 | 0 | 0 | 1 | 1 | 0 | 0 | 1 | 1 | 1 | 1 | 1 | 0 | 0 | 1 | 1 | 1 | 1 | 0 | 1 | 1 | 1 | 1 | 25 |
| Guisso, 2018 | 1 | 1 | 1 | 1 | 1 | 1 | 0 | 1 | 1 | 1 | 1 | 1 | 0 | 0 | 0 | 0 | 1 | 1 | 0 | 0 | 1 | 1 | 1 | 1 | 0 | 0 | 0 | 1 | 1 | 1 | 1 | 0 | 1 | 1 | 1 | 1 | 25 |
| Oommen, 2018 | 1 | 0 | 1 | 1 | 1 | 1 | 1 | 1 | 1 | 0 | 1 | 0 | 0 | 0 | 0 | 0 | 0 | 1 | 0 | 0 | 1 | 1 | 1 | 1 | 1 | 0 | 0 | 0 | 1 | 1 | 1 | 0 | 1 | 1 | 1 | 1 | 22 |
| Malek, 2019 | 1 | 1 | 1 | 1 | 1 | 1 | 1 | 1 | 1 | 1 | 1 | 0 | 0 | 0 | 0 | 0 | 1 | 1 | 0 | 0 | 1 | 1 | 0 | 0 | 0 | 0 | 0 | 1 | 1 | 1 | 1 | 0 | 1 | 1 | 1 | 1 | 23 |
| Sadek, 2019 | 1 | 0 | 1 | 1 | 1 | 1 | 1 | 1 | 1 | 1 | 1 | 1 | 1 | 0 | 0 | 0 | 0 | 1 | 0 | 1 | 1 | 1 | 1 | 1 | 1 | 0 | 0 | 0 | 1 | 1 | 0 | 0 | 1 | 1 | 1 | 1 | 25 |
| Al-Zalabani, 2019 | 1 | 0 | 1 | 1 | 1 | 1 | 1 | 1 | 1 | 1 | 1 | 1 | 0 | 0 | 0 | 0 | 0 | 1 | 0 | 1 | 1 | 1 | 1 | 1 | 1 | 0 | 0 | 1 | 1 | 1 | 1 | 0 | 1 | 1 | 1 | 1 | 26 |
| Jenabi, 2020 | 1 | 1 | 1 | 1 | 1 | 1 | 1 | 1 | 1 | 1 | 1 | 1 | 1 | 0 | 0 | 0 | 1 | 1 | 0 | 1 | 1 | 1 | 0 | 0 | 0 | 0 | 0 | 1 | 0 | 1 | 1 | 0 | 1 | 1 | 1 | 1 | 25 |
| Gerges, 2020 | 1 | 0 | 1 | 1 | 1 | 1 | 1 | 1 | 1 | 1 | 1 | 1 | 0 | 0 | 0 | 0 | 0 | 1 | 0 | 1 | 1 | 1 | 0 | 1 | 1 | 0 | 0 | 1 | 1 | 1 | 1 | 0 | 1 | 1 | 1 | 1 | 25 |
| Shamsedine, 2020 | 1 | 1 | 1 | 1 | 1 | 1 | 1 | 1 | 1 | 1 | 1 | 1 | 1 | 1 | 0 | 0 | 1 | 1 | 0 | 1 | 1 | 1 | 1 | 1 | 1 | 0 | 0 | 1 | 1 | 1 | 1 | 0 | 1 | 1 | 1 | 1 | 30 |
| Alkhalidy, 2021 | 1 | 0 | 1 | 1 | 1 | 1 | 0 | 1 | 1 | 1 | 1 | 1 | 0 | 0 | 0 | 0 | 0 | 1 | 0 | 0 | 1 | 1 | 1 | 1 | 0 | 0 | 0 | 1 | 1 | 0 | 0 | 0 | 1 | 1 | 1 | 1 | 21 |
| Kheirouri, 2022 | 1 | 0 | 1 | 1 | 1 | 1 | 1 | 1 | 1 | 1 | 1 | 1 | 0 | 0 | 0 | 0 | 0 | 1 | 0 | 1 | 1 | 1 | 1 | 1 | 1 | 0 | 0 | 1 | 1 | 1 | 1 | 0 | 1 | 1 | 1 | 1 | 26 |
| Arafa, 2022 | 0 | 0 | 1 | 1 | 1 | 1 | 0 | 1 | 1 | 1 | 1 | 0 | 1 | 0 | 0 | 0 | 0 | 1 | 0 | 0 | 1 | 1 | 1 | 1 | 1 | 0 | 0 | 1 | 1 | 1 | 1 | 0 | 1 | 1 | 1 | 1 | 23 |
| Hajj, 2022 | 1 | 0 | 1 | 1 | 1 | 1 | 1 | 1 | 1 | 1 | 1 | 1 | 1 | 0 | 0 | 0 | 0 | 1 | 0 | 1 | 1 | 1 | 1 | 1 | 0 | 0 | 0 | 1 | 1 | 1 | 1 | 0 | 1 | 1 | 1 | 1 | 26 |
| Burhani, 2023 | 0 | 1 | 1 | 1 | 1 | 1 | 0 | 1 | 1 | 1 | 1 | 1 | 0 | 0 | 0 | 0 | 1 | 1 | 0 | 1 | 1 | 1 | 1 | 1 | 1 | 0 | 0 | 1 | 1 | 1 | 1 | 0 | 1 | 1 | 1 | 1 | 26 |
| Alshaban, 2023 | 0 | 0 | 1 | 1 | 1 | 1 | 0 | 1 | 1 | 1 | 1 | 1 | 0 | 0 | 0 | 0 | 0 | 1 | 0 | 1 | 1 | 1 | 1 | 1 | 0 | 0 | 0 | 1 | 1 | 1 | 1 | 0 | 1 | 1 | 1 | 1 | 23 |
| Al-Mamari, 2021 | 1 | 1 | 1 | 1 | 1 | 1 | 1 | 1 | 1 | 1 | 1 | 1 | 0 | 0 | 0 | 0 | 0 | 1 | 0 | 1 | 1 | 1 | 1 | 1 | 1 | 0 | 0 | 1 | 1 | 1 | 1 | 0 | 1 | 1 | 1 | 1 | 27 |

**Supplementary Tables**

Supplementary Table S8. Summary of Egger's *p* Values for the analyzed risk factors.

| Risk Factor | Egger's *p* Value |
|---|---|
| Cesarean Delivery | 0.046 |
| Consanguinity | 0.265 |
| Family History | 0.738 |
| Male Gender | 0.944 |
| Maternal Age ≥35 Years | 0.277 |

### Supplementary Figures

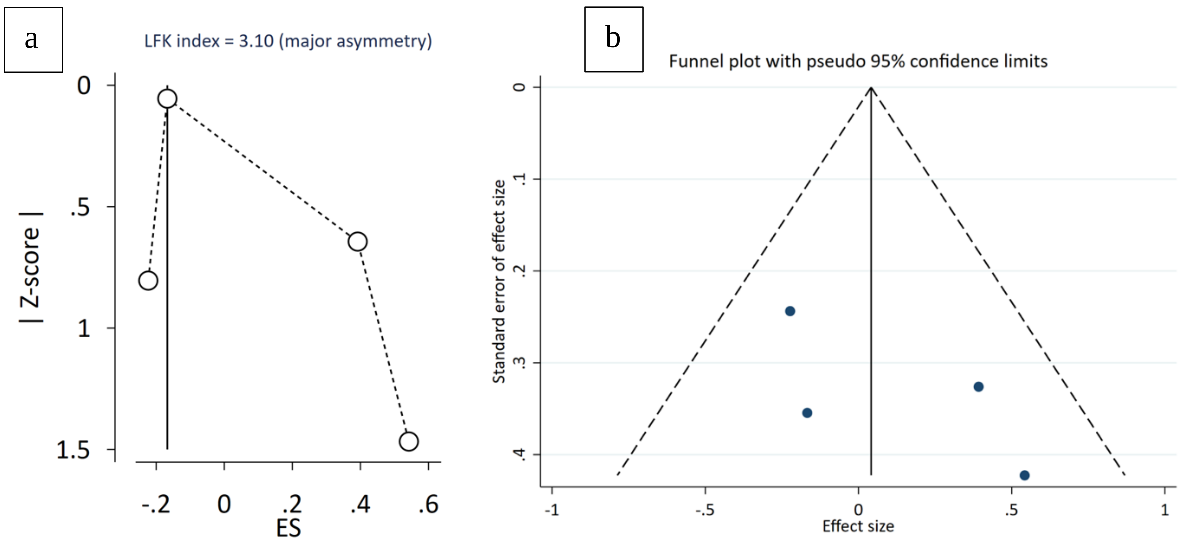

Supplementary Figure S1. (a) Doi Plot and LFK index for Maternal Age ≥35 Years; (b) Funnel Plot for Maternal Age ≥35 Years.

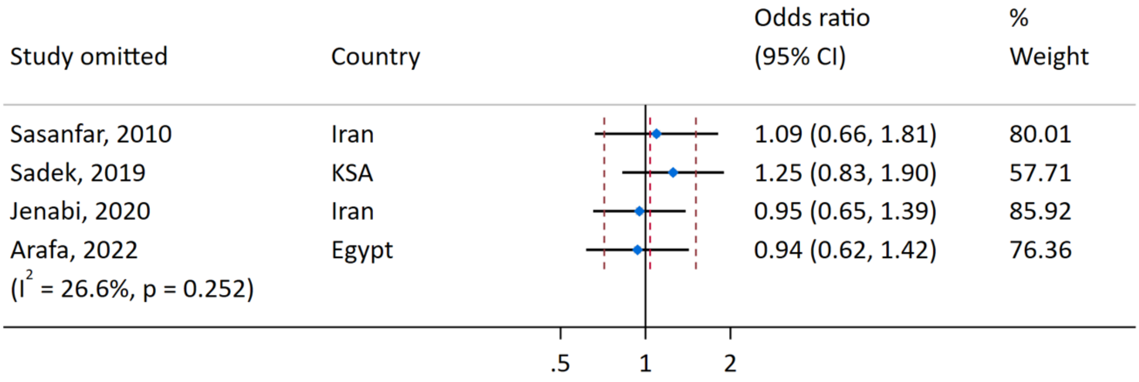

Supplementary Figure S2. Leave-one-out analysis for Maternal Age ≥35 Years.

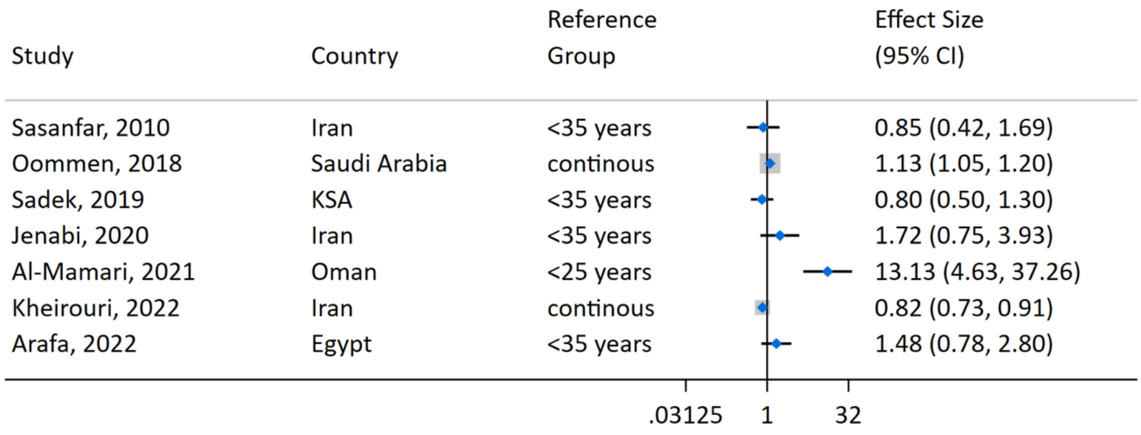

Supplementary Figure S3. Forest Plot for Maternal Age (not pooled).

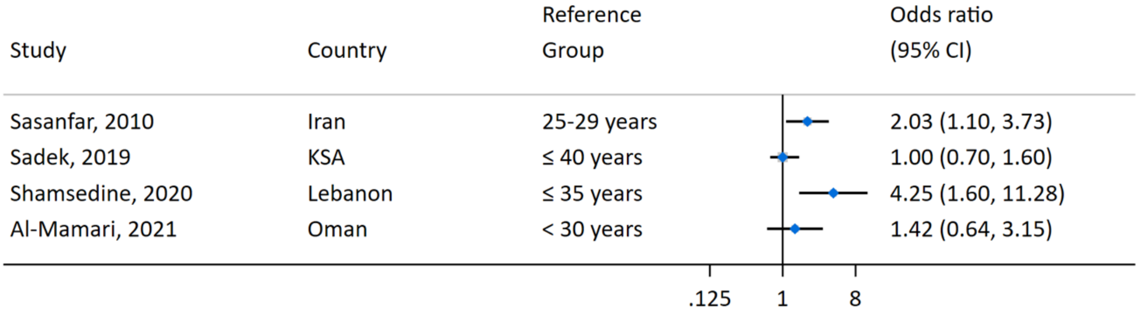

Supplementary Figure S4. Forest Plot for Paternal Age (not pooled).

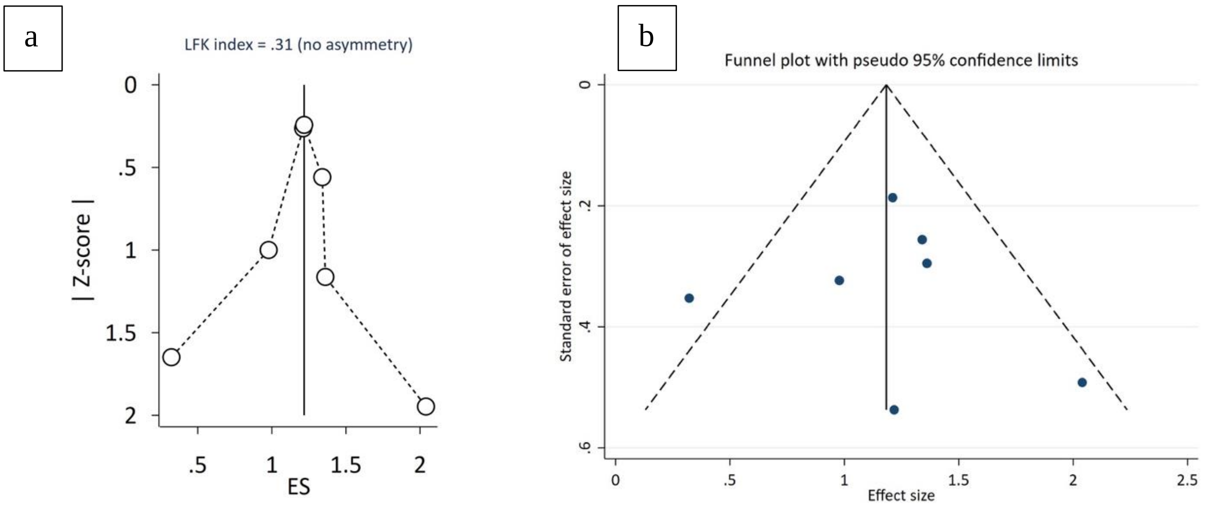

Supplementary Figure S5. (a) Doi Plot and LFK index for Male Gender; (b) Funnel Plot for Male Gender.

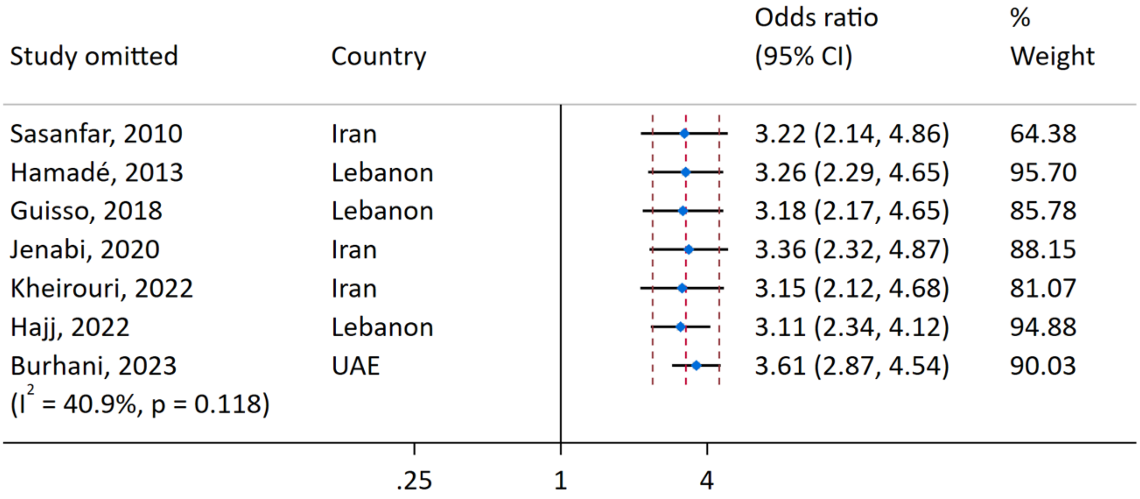

Supplementary Figure S6. Leave-one-out analysis for Male Gender.

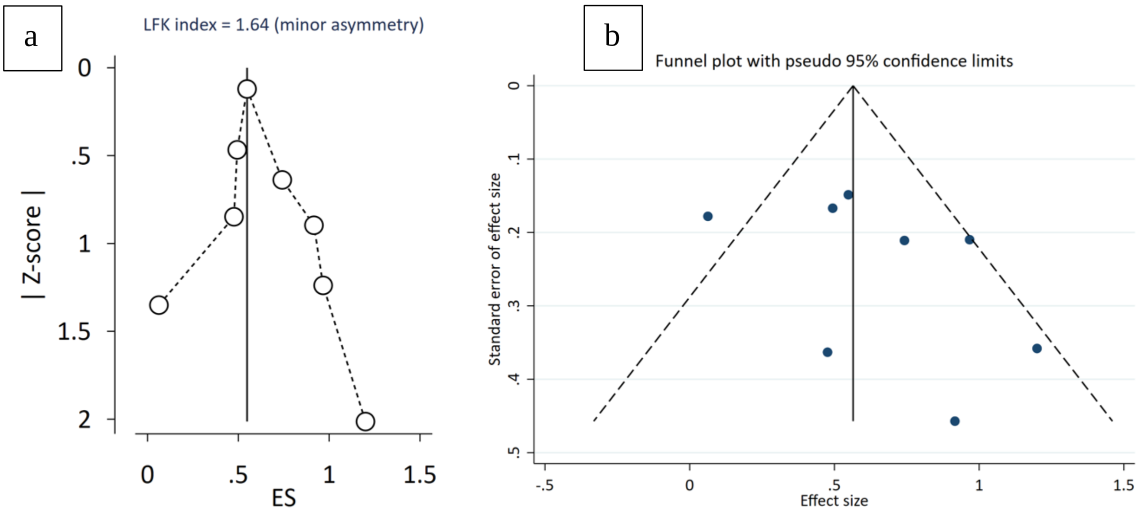

Supplementary Figure S7. (a) Doi Plot and LFK index for Consanguinity; (b) Funnel Plot for Consanguinity.

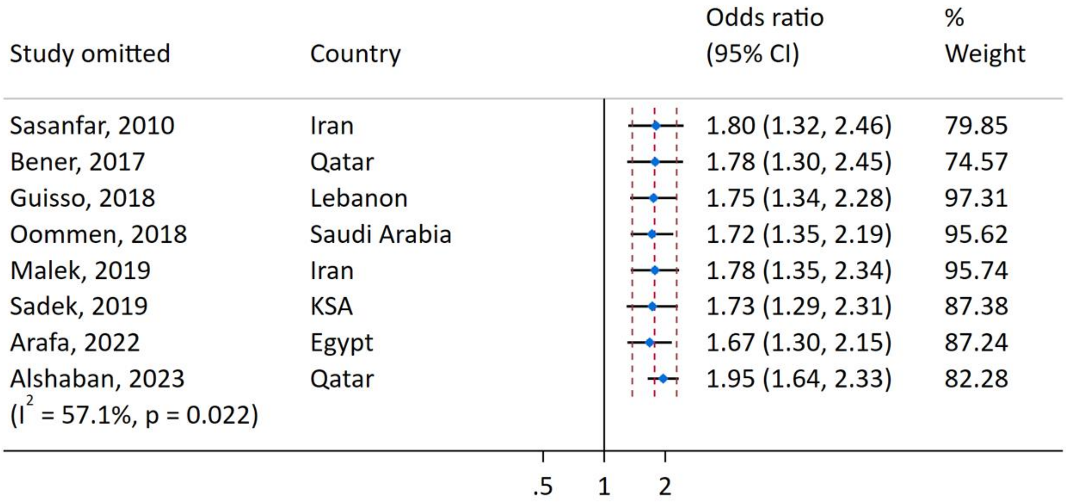

Supplementary Figure S8. Leave-one-out analysis for Consanguinity.

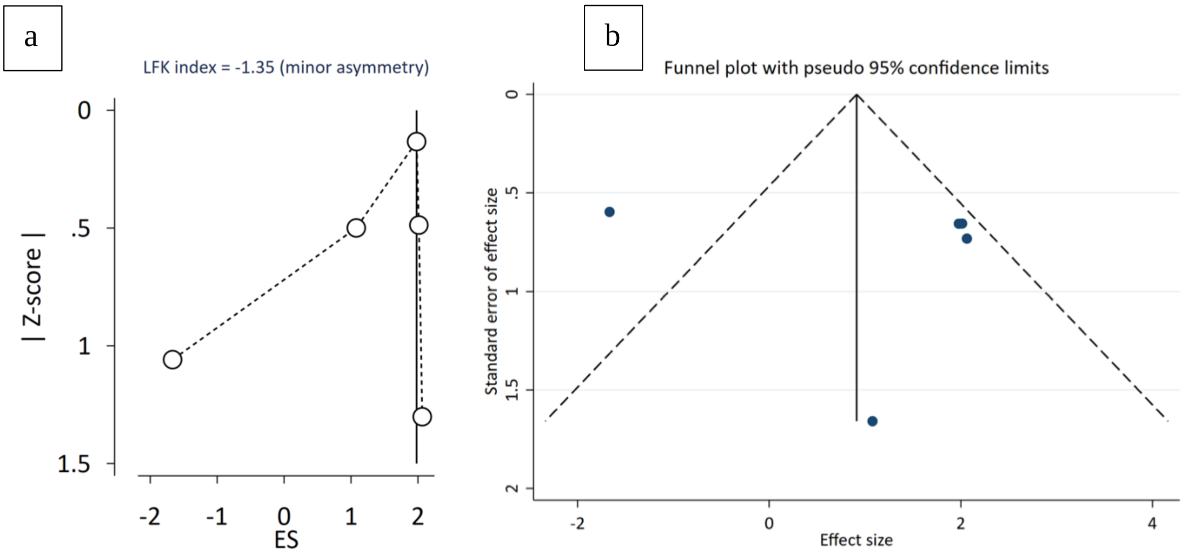

Supplementary Figure S9. (a) Doi Plot and LFK index for Family History of ASD; (b) Funnel Plot for Family History of ASD.

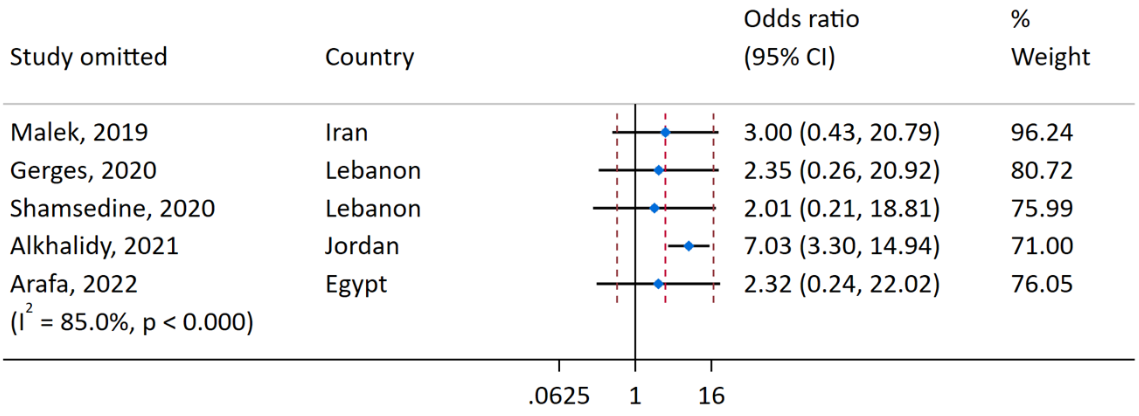

Supplementary Figure S10. Leave-one-out analysis for Family History of ASD.

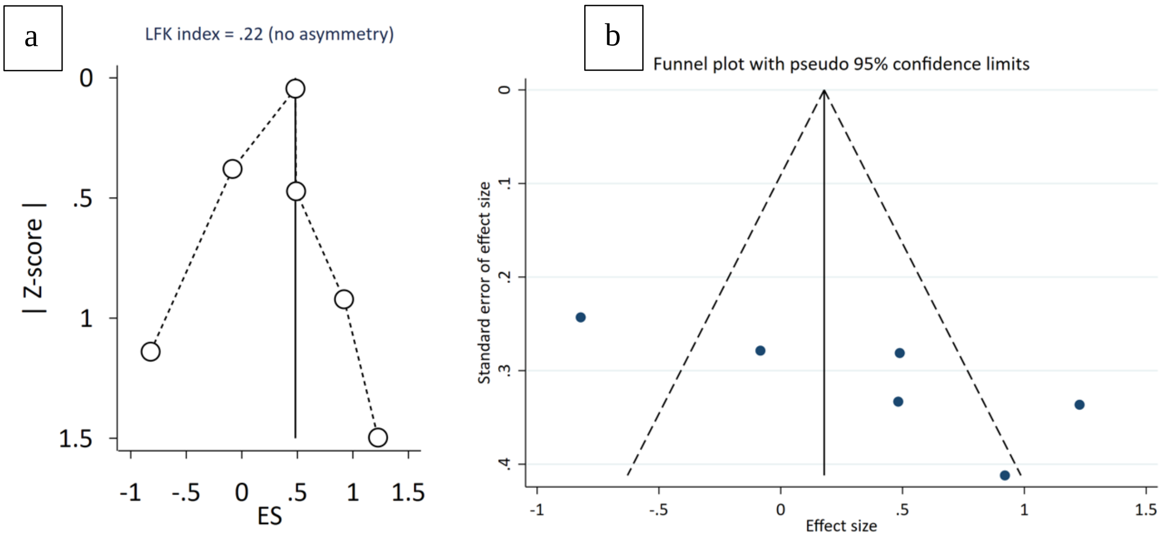

Supplementary Figure S11. (a) Doi Plot and LFK index for Cesarean Delivery; (b) Funnel Plot for Cesarean Delivery.

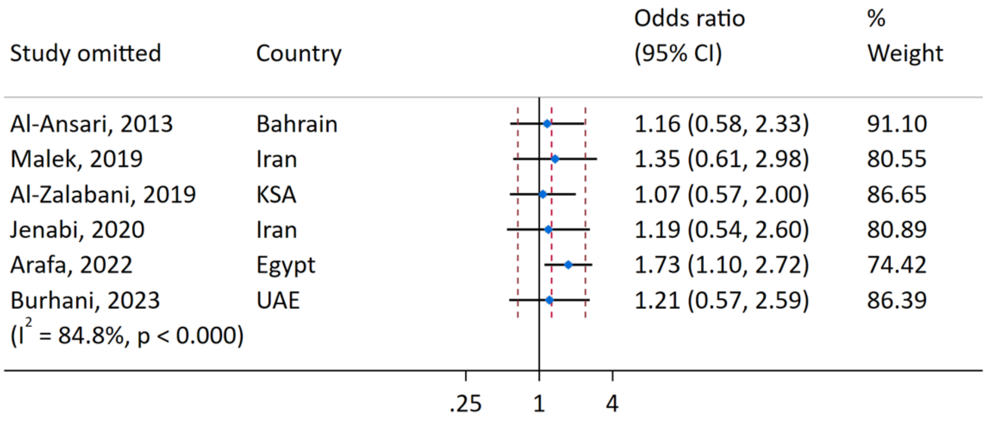

Supplementary Figure S12. Leave-one-out analysis for Cesarean Delivery.

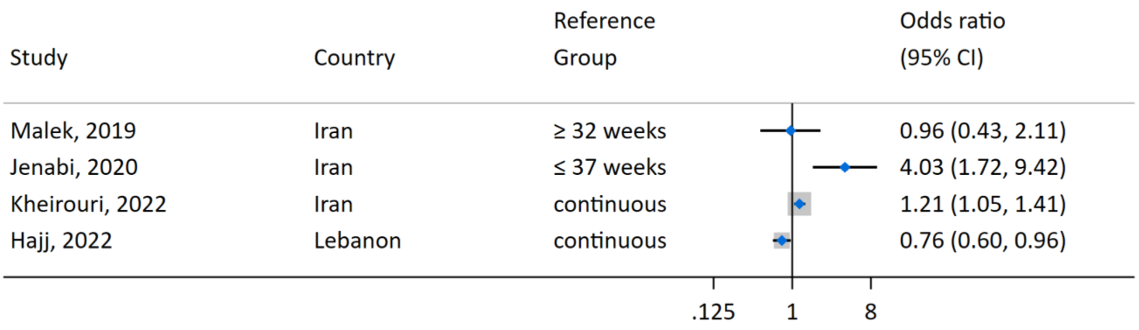

Supplementary Figure S13. Forest Plot for Gestational Age (not pooled).
